# Circulating thiamine diphosphate, dietary intake, and transketolase function are non-equivalent measures of thiamine status

**DOI:** 10.64898/2026.09.12.26362925

**Authors:** Katie A. Edwards, Nannette Cowen, Patricia C. Wolfe, Michaela Lynch

## Abstract

We conducted a pilot study in adults with and without type 2 diabetes (T2D), measuring whole-blood thiamine diphosphate (TDP) concentrations alongside erythrocyte transketolase activity (TKT) and activity coefficients (ETKAC), and evaluated these findings alongside dietary and ETKAC data from the U.K. National Diet and Nutrition Survey (NDNS). In the pilot cohort, circulating TDP concentrations did not differ between T2D and control groups, and ETKACs were largely sufficient, yet absolute TKT activities varied approximately sixfold among participants. Magnesium supplementation of erythrocyte lysates increased TKT activity, demonstrating that cofactor availability beyond TDP can influence functional measurements. Discordance between circulating TDP and functional status was also observed, including ETKAC-defined deficiency despite a TDP concentration within the reference range. Among participants with T2D, TDP was positively associated with fasting glucose and HbA1c, a relationship absent in controls. In NDNS analyses, higher thiamine intake and dietary density were strongly associated with lower ETKAC-defined non-sufficiency, although substantial non-sufficiency persisted despite adequate intake. Dietary thiamine density, ethnicity, age, and magnesium intake were independently associated with ETKAC status. Collectively, these findings support the conclusion that circulating TDP, dietary intake, and ETKAC provide related, but non-equivalent measures and support integrating both direct and functional assessments of thiamine status.

**Graphical Abstract:** **Caption:** Dietary thiamine intake, circulating TDP, and transketolase function are related but non-equivalent measures of thiamine status.

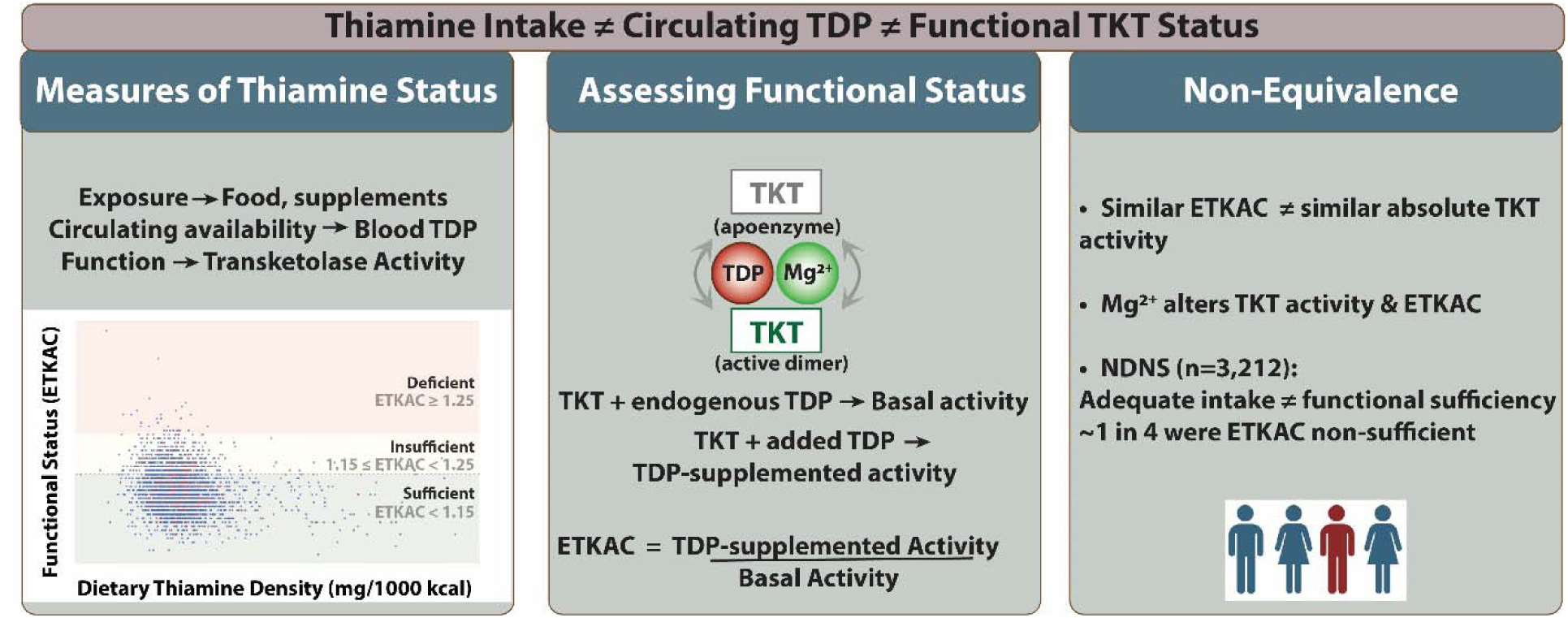

**Highlights:**

- Thiamine-TKT relationships differ between T2D and control participants
- TKT activities varied as much as six-fold participant to participant
- ETKAC ratio outputs mask absolute values of TKT activity
- TKT activity increases with magnesium supplementation ex vivo
- ETKAC-defined thiamine non-sufficiency persists despite adequate reported intake

## Introduction

In 2023, approximately 12% of the U.S. population (∼40.1 million people) was estimated to have diabetes, including 11 million adults with undiagnosed diabetes, while another 115.2 million U.S. adults had prediabetes, a major risk state for Type 2 diabetes (T2D).^1^ Approximately 1.5 million Americans were newly diagnosed with diabetes in 2023,^1^ with T2D accounting for 90-95% of diagnosed diabetes cases. Projections suggest that the number of diagnosed diabetes cases will increase to 60.6 million people by 2060 in the U.S. alone.^2^

T2D is a complex metabolic disorder characterized by impaired glucose homeostasis arising from the combined effects of insulin resistance and progressive β-cell dysfunction.^3^ These defects arise from interactions among genetic susceptibility, lifestyle, and environmental factors and are accompanied by broader disturbances in intermediary metabolism, including altered lipid and protein handling.^4–6^ Importantly, these systemic changes reflect disruptions in enzyme-dependent metabolic pathways that govern cellular energy balance and redox status.^6^ Many of these enzymes rely on vitamins as cofactors or precursors of essential substrates, providing a mechanistic context for examining nutrient-dependent enzyme function in diabetes, particularly within pathways that link glucose metabolism to cellular redox regulation.

Glucose metabolism is distributed across connected pathways that coordinate energy production and biosynthesis. Following its phosphorylation to glucose-6-phosphate, glucose can enter the pentose phosphate pathway (PPP), which generates NADPH for cellular antioxidant defense and supports redox homeostasis, as well as ribose-5-phosphate (R5P) for nucleotide synthesis.^7,8^ Alternatively, glycolytic flux yields pyruvate, which enters the tricarboxylic acid (TCA) cycle, producing reducing equivalents (NADH and FADH_2_) that drive oxidative phosphorylation and ATP generation. Under conditions where mitochondrial oxidative metabolism is limited, however, pyruvate may instead be reduced to lactate to regenerate NAD+, thereby sustaining glycolytic ATP production. The regulation of the flux through these pathways is highly dynamic,^8,9^ and influenced by factors relevant to diabetes, including hyperglycemia, insulin signaling, and oxygen availability.^6,10–12^

Central metabolic pathways involved in glucose utilization, including the TCA cycle and the PPP, require thiamine (vitamin B1) as a cofactor for key enzymes. In its active form, thiamine diphosphate (TDP) is required for pyruvate dehydrogenase, which links glycolysis to the TCA cycle, and transketolase (TKT), which regulates carbon flux in the PPP. It further serves as a cofactor for α-ketoglutarate dehydrogenase in the TCA cycle, further integrating thiamine status with oxidative metabolism, and branched-chain α-ketoacid dehydrogenase in branched chain amino acid catabolism. Through these roles, thiamine availability can influence both energy production and cellular redox balance. Clinical and experimental studies have examined the relationship between thiamine status and diabetes, including investigations of thiamine supplementation and thiamine analogs.^12–16^ While some studies suggest improvement in glycemic control or markers of metabolic stress, findings have been inconsistent, and mechanistic interpretation remains limited.^16–19^

Thiamine status is typically assessed in populations at risk for thiamine deficiency, historically focusing on regions with limited dietary diversity and heavy reliance on white rice.^20,21^ Although food fortification has reduced the prevalence of overt deficiency in developed countries, inadequate intake, malabsorption, and increased metabolic demand remain clinically relevant contributors.^22–27^ Manifestations of thiamine deficiency typically yield cardiac and nervous system symptoms with greater risks of fatality to infants.^22,23^ Conditions such as alcohol use disorder,^24^ obesity^25,26^, bariatric surgeries,^25^ and hyperemesis gravidarum^27^ have all been associated with impaired thiamine status.

Despite growing interest in thiamine status in metabolic disease,^12,16,28–30^ thiamine deficiency likely remains underrecognized due to nonspecific clinical manifestations and limited routine testing.^31^ Large-scale population datasets often rely on dietary intake estimates rather than direct quantification of thiamine by chromatography or enzymatic functional assessment.^32,33^ However, the U.K. National Diet and Nutrition Survey (NDNS) includes data from the erythrocyte transketolase activity (ETKA) assay, which provides insight into cofactor availability at the enzyme level. ETKA assays quantify the increase in TKT activity following TDP supplementation, with larger increases indicating reduced baseline cofactor saturation, typically expressed as the ETKA coefficient (ETKAC). However, the interpretation of ETKA results remains complex, as enzyme activity may be influenced not only by thiamine availability but also by factors that affect enzyme structure, cofactor binding, and the cellular environment.^34,35^ Furthermore, population datasets and published studies frequently report either ETKAC measurements or direct thiamine/TDP concentrations without both measures being available in the same samples (Table 1).^20,36–38^

**Table 1.** Thiamine status measures available across data sources.

| Data Source | Population | Dietary intake | ETKA | Thiamine/TDP levels |
| --- | --- | --- | --- | --- |
| Internal pilot study | Adults (18-65) | No | Yes | Yes |
| CDC NHANES | Adults (18-65) | Yes | No | No |
| UK NDNS | Adults (18-65) | Yes | Yes | No |

To address this gap, we conducted a pilot study with participants with and without T2D, integrating direct measurements of TDP with functional assessment of TKT activity, including basal and TDP-stimulated activity, as well as ETKAC determinations. These data were evaluated alongside glycemic markers and compared to patterns observed in the NDNS to better understand thiamine status at both the biochemical and functional levels. Our findings reveal substantial variability in functional enzyme capacity in adults aged 18-65 and highlight the influence of magnesium-dependent cofactor interactions on TKT activity, with implications for interpreting thiamine status in metabolic disease.

## Materials and Methods

NADH, TDP, D-ribose-5-phosphate, hemoglobin, and α-glycerophosphate dehydrogenase-triosephosphate isomerase (α-GDH-TPI) from rabbit muscle were purchased from Sigma. Drabkin’s Reagent was purchased from Ricca Chemical Company. Ficoll-Paque Plus was purchased from Cytiva. All other supplies were purchased from VWR or Fisher Scientific.

### Participant recruitment and body measurements

We recruited adult participants aged 18-65 with Type 2 Diabetes (T2D) or controls. The study was approved under Binghamton University IRB protocol # STUDY00003752. The inclusion criteria were healthy, prediabetic, or type 2 diabetic; able to tolerate a fingerstick and venous blood draw with no history of fainting or dizziness associated with procedures; and living, working, or attending school in the local Binghamton, NY, area. Exclusion criteria were history of significant cardiovascular, pulmonary, hepatic, renal, hematological, metabolic, or gastrointestinal disease; lost more than 20 lbs. in the past 30 days, having had a gastric bypass, lap band placement, or any GI surgery; donated blood and/or platelets, white blood cells, or plasma within the past 60 days; have received any of the following diagnoses; anemia, celiac disease, IBD, currently pregnant, or less than 6 weeks postpartum, cancer within the last 5 years, alcohol/drug/chemical dependence within the last year, eating disorder (anorexia nervosa, bulimia) in the past 5 years; or adherence to a strict vegan diet. Participants gave informed consent and were asked to fast for 12 hours before the appointment with Dr. Cowen (Nurse Practitioner). At this appointment, body measurements (height, weight, BMI, waist circumference, blood pressure and pulse) were collected, and POC glucose and HbA1c assays were carried out by the respective manufacturer’s instructions. A Contour Next EZ Blood glucose monitoring system with Contour Next Blood Glucose test strips and lancets was used to assess fasting blood glucose. A1CNow Self Check device (analyzer, lancet, blood collector was used to assess HbA1c. Venous blood was collected at United Health Services (UHS) Lab Services in Vestal, NY, in lavender-top EDTA tubes for transketolase and TDP assays and hemoglobin analysis. TDP measurements were carried out on 4 mL of whole blood submitted to the Mayo Clinic Laboratories for LC-MS^2^ analysis, ordered under CPT code 84425 by UHS Lab Services. The TDP concentrations for two control participants were unavailable due to a logistical issue between sample collection and analysis.

### Isolation of erythrocytes

Blood samples were received and processed within 30 minutes after they were drawn. Ficoll-Paque separation: In a 15 mL centrifuge tube, 2 mL Balanced Salt Solution (BSS) was freshly prepared by combining 0.2 mL Solution A (0.1% anhydrous D-glucose, 50 μM CaCl_2_ x 2H_2_O, 980 μM MgCl_2_ x 6H_2_O, 5.4 mM KCl, 145 mM Tris) with 1.8 mL 0.819% sodium chloride. 2 mL of the fresh whole blood sample was added to the BSS and mixed. In a separate 15 mL centrifuge tube, 3 mL of Ficoll-Paque PLUS (FPP) was added and the whole blood/BSS mixture was carefully layered on top. The tube was then centrifuged at 400xg and 18°C for 35 min. After centrifugation, the plasma layer was removed using a plastic Pasteur pipette. The erythrocytes were washed 3X by adding regular saline at 2X the volume of erythrocytes, centrifuging at 400x*g* and 4°C for 10 min., and removing the saline layer with a Pasteur pipette. The washed erythrocytes were stored at −80°C until analysis.

### Erythrocyte transketolase assays

The extraction and assay protocol for transketolase activity was adapted from Jones et al.^37,39^ Briefly, 25 μL of washed erythrocytes were diluted with 50 μL water and allowed to stand for 10 min., before centrifuging for 10 min. at 4°C and 4,000*xg*. The hemolysate was diluted 10-fold in 100 mM Tris, pH 7.6 and thoroughly mixed. 15 μL of either Tris buffer alone or 3.6 mg/mL TDP in Tris buffer was added to triplicate columns six wells of a UV transparent plate (Greiner). 30 μL of diluted supernatants or control were added to triplicate wells, with and without TDP. HPLC grade water diluted identically in Tris buffer was used as a control and treated the same way as the standards. The plate was shaken at 600 rpm for 30 seconds, sealed, then incubated at 37°C for 15 min. During the incubation, a mixture of NADH (256 μM), D-ribose-5-phosphate (16.26 mM), and α-GDH-TPI (0.208 U/mL) was prepared in Tris buffer containing 0.004% (w/v) Tween-20. 200 μL of this solution was added to all wells of the plate, which was then immediately transferred to a plate reader (Varioskan, ThermoFisher) set at 37°C and read at 340 nm every minute over 60 min. to monitor NADH depletion with 5 seconds shaking at 600 rpm before each read. A subset of six samples was analyzed by comparing the basal and TDP-supplemented TKT rates in the presence or absence of added MgCl_2_ spiked into the 15 μL volume prior to incubation for 15 min. at 37°C.^34^ The data were analyzed following the process reported by Jones et al.^37^ The ETKAC values were reported as the ratio between TDP-supplemented and basal activity. The specific activity of the sample was determined by taking the basal and supplemented rates and dividing by the micrograms of hemoglobin present in the assay well. The hemoglobin content of hemolysates (10 μL) was determined in triplicate at 540 nm against freshly prepared standards of human hemoglobin (Sigma) and 200 μL of the Drabkin’s reagent in a clear Corning Costar microplate.

### NDNS Survey results

Data from the UK National Diet and Nutrition Survey (NDNS; 2008–2019, Years 1–11)^40^ were analyzed to assess the prevalence of functional thiamine insufficiency, defined by elevated erythrocyte transketolase activation coefficient (ETKAC), and its relationship to dietary thiamine intake. Publicly available datasets were downloaded from the UK Data Service website and combined across survey periods (years 1-4 (2008-2012), years 5-6 (2013-2014), years 7-8 (2015-2016), and years 9-11 (2017-2019)). In this study, only participants who completed a food diary over three to four days were invited to undergo physical body measurements and submit a blood sample. Information on study design, nutrient composition, and blood sample handling is available in the study documentation.

Our interest was not in reporting the overall prevalence of thiamine deficiency in the U.K. population and the contributing factors at the population level, but rather in how parameters relevant to our pilot study affect ETKAC status on an individual basis. As such, unweighted analyses were used to assess relationships between ETKAC and dietary intake variables to preserve underlying distributional characteristics and avoid possible instability introduced by weighting within smaller subgroups (e.g., diabetics, ETKAC-deficient).

Data downloaded from the UK Data Service website were processed using Stata (Version 18) or JMP (Version 17). As the questionnaire structure varied slightly across survey years, a unified variable representing diabetes status was generated. Participants were classified as diabetic based on affirmative responses to survey items indicating a diagnosis of diabetes or diabetes-related fasting blood participation (Years 1-3 and 11), or direct diabetes status variables (Years 4-10). We also assessed diabetes status based on biochemical parameters. Fasting plasma glucose was grouped into <5.6 mmol/L, 5.6-<7.0 mmol/L, and ≥7.0 mmol/L as normal, prediabetic, and diabetic, respectively. HbA1c was grouped into <5.7%, 5.7-<6.5%, and ≥6.5% as normal, prediabetic, and diabetic, respectively. These ranges correspond to ADA-defined normal, prediabetic, and diabetic categories.^41^

Across all survey years, 15,655 individuals were included, of whom 5,657 had available ETKAC measurements and information on thiamine intake, including 3,212 adults aged 18 to 65. Within this subgroup, 2,817 were classified as non-diabetic and 131 self-reported as diabetic. We analyzed data from this population using unweighted analyses, as we were interested in the relationship between dietary thiamine intake and ETKAC values at the individual level. We used data on total thiamine intake (including supplements) in these analyses, both on a mg/day basis and mg/1000 kcal basis. In other individual comparisons, we assessed ETKAC values in relation to fasting blood sugar, %HbA1c, total magnesium intake (including supplements), total sugar intake, energy intake, BMI, age, sex, and ethnicity.

### Statistical analysis

For analysis of the pilot data, linear regression was used to evaluate associations between continuous variables, and slopes were compared using the extra sum-of-squares *F* test in GraphPad (Version 11.1.0). Comparisons of measured parameters between control and T2D groups (Table 2) were carried out using unpaired t-tests with Welch’s correction. The effects of added Mg^2+^ on TKT activity were carried out using paired t-tests since measurements were carried out on the same samples under +/-Mg^2+^ conditions.

**Table 2.** Participant characteristics and measured parameters.

| Parameter | Control | Type 2 Diabetics | t-test p-value |
| --- | --- | --- | --- |
| *Patient population and clinical measurements* |  |  |  |
| N | 18 | 6 |  |
| Sex (F/M) | 15/3 | 4/2 |  |
| Age | 38.6±2.7 | 50.5±7.1 | 0.166 |
| BMI | 25.8±1.1 | 27.7±1.9 | 0.412 |
| *Blood measurements* |  |  |  |
| Fasting blood glucose (mg/dL) | 92.9±1.94<br>(67-105) | 119.7±12.7<br>(76-159) | 0.089 |
| % HbA1c | 5.0±0.1<br>(4.2-5.8) | 6.3±0.36<br>(5.3-7.5) | 0.016* |
| Hemolysate Hb (g/dL) | 12.34±0.16<br>(11.3-13.3) | 11.84±0.32<br>(11.0-13.2) | 0.202 |
| [TDP] (nM)<br>(range) | 133.3±9.9 (n=16*)<br>(72-218) | 125.2±12.5<br>(83-170) | 0.618 |
| Basal TKT activity (pmol/min) | 120.7±9.6<br>(40.6-212.3) | 139.6±18.9<br>(80.8-213.7) | 0.417 |
| Supplemented TKT activity (pmol/min) | 125.6±11.85<br>(39.6-212.1) | 149.5±30.9<br>(91.2-222.7) | 0.317 |
| ETKAC | 1.040±0.015<br>(0.913-1.142) | 1.085±0.0436<br>(0.968-1.275) | 0.383 |
| Basal TKT activity (pmol/min/μg Hb) | 0.324±0.030<br>(0.114-0.533) | 0.398±0.060<br>(0.204-0.615) | 0.289 |
| Supplemented TKT activity (pmol/min/μg Hb) | 0.337±0.030<br>(0.104-0.533) | 0.431±0.065<br>(0.230-0.641) | 0.230 |

Statistical analyses for NDNS data were carried out using Stata (Version 18). Functional thiamine status was assessed using ETKAC thresholds of ≥1.15 and ≥1.25 to indicate insufficiency and deficiency, respectively. In most analyses (as specified within), ETKAC data for insufficiency (n=758) and deficiency (n=42) were pooled into a non-sufficiency category (ETKAC ≥1.15, n=800) for comparison with sufficiency (n=2,412) to address the low deficiency sample number. Thiamine intake was grouped based on <1 mg/day, 1-<2 mg/day, 2-<3 mg/day, and ≥3 mg/day, including supplements. Thiamine intake normalized to 1000 kcal (termed thiamine density within) was grouped based on <0.5 mg/1000 kcal, 0.5 -<0.75 mg/1000 kcal, 0.75 - <1.0 mg/1000 kcal, and ≥1 mg/1000 kcal, including supplements. An RDA for thiamine of approximately 1.0 mg/day or based on the caloric intake of 0.5 mg/1000 kcal has been historically used,^1,26,42,43^ while the other bins were investigator-defined. Thiamine supplement users were grouped based on ≤1.5 mg/day or >1.5 mg/day to delineate between the amounts found in standard multivitamin formulations (≤1.5 mg/day), relative to high dose B complex formulations, or thiamine only formulations (12.5-500 mg). Magnesium intake was grouped based on <200 mg/day, 200-<300 mg/day, 300-<400 mg/day, and ≥400 mg/day for descriptive and categorical analyses and modeled continuously, scaled per 100 mg/day, in multivariable analyses.

Associations between categorical variables and functional thiamine status were evaluated using Pearson’s chi-squared tests. Continuous variables were compared between ETKAC-defined sufficient and non-sufficient groups using Wilcoxon rank-sum tests. Multivariable logistic regression was used to identify factors independently associated with functional non-sufficiency (ETKAC ≥1.15). The primary model included thiamine density, magnesium intake, age, sex, BMI, ethnicity, and fasting blood glucose. Adjusted odds ratios (ORs) and 95% confidence intervals (CIs) are reported. HbA1c was evaluated in a separate model in place of fasting glucose. Age was modeled as a continuous variable and scaled in 10-year increments to facilitate interpretation of odds ratios. Overall significance of categorical predictors in the multivariable model was assessed using omnibus Wald tests. The independent contribution of individual predictors to model fit was further evaluated using likelihood-ratio tests comparing the full model with otherwise identical nested models omitting each predictor individually. Model-adjusted predicted probabilities and 95% CIs were estimated following logistic regression, with pairwise comparisons used where indicated. Continuous data are presented as median (IQR) for NDNS analyses and mean±SEM for the pilot study, unless otherwise indicated. All tests were two-sided and statistical significance was defined as p<0.05. Multiple linear regression was additionally used to evaluate the TDP-glycemic measure associations after adjustment for age.

## Results

### Pilot study results

The characteristics of the participants in our pilot study are presented in Table 2, along with measured parameters (thiamine diphosphate levels, blood glucose, % glycated hemoglobin (HbA1c), basal and TDP-activated transketolase specific activity, and ETKACs). Our local recruitment yielded 6 participants with a clinical diagnosis of T2D and 18 controls, all aged 18-65.

Values presented represent the average and SEM. The ranges are presented in parentheses. An unpaired two-tailed t-test with Welch’s correction was run to determine the significance (assessed at p≤0.05) of differences between control and T2D participants.

*Two TDP values from control participants were unavailable due to a delay in processing.

In this population, the hemolysate hemoglobin concentration did not differ significantly, but HbA1c values were significantly higher in T2D participants than in controls (Table 2). Fasting blood glucose was higher for T2D participants, but this difference did not reach statistical significance. There were no significant differences in hemoglobin concentration, TDP concentration, basal or supplemented TKT activity, or ETKAC between T2D and control participants. Across the observed range of whole blood TDP concentrations, most participants fell within the established reference range (70-180 nM). One participant was near the lower end of the TDP reference range (72 nM), while another was near the upper end (179 nM), and two exceeded it (185 and 218 nM).

### Thiamine supplementation and TDP concentrations

Five participants reported regular use of thiamine-containing supplements (0.8 to 25 mg/day). Whole blood TDP concentrations in these individuals (170-218 nM) clustered at the upper end of or above the reference range, whereas concentrations were generally lower among participants not reporting regular thiamine supplementation (Fig. S1). Despite the wide range of reported supplemental doses, TDP concentrations among supplement users were relatively similar. This is consistent with the saturable absorption and cellular uptake of thiamine.^44,45^

### Erythrocyte transketolase activity coefficients (ETKAC)

We then considered transketolase activity as a measure of biological activity, including ETKAC and the specific activities from both basal and supplemented conditions. An increase above 15% over basal activity levels with TDP supplementation are commonly interpreted as thiamine insufficiency, while an increase above 25% indicates deficiency.^46–48^ With one exception, when looking at the ratio data, all participants were in a sufficient range (ratio <1.15) irrespective of their circulating TDP concentrations (Table 2, Fig. 1).

**Fig. 1.**
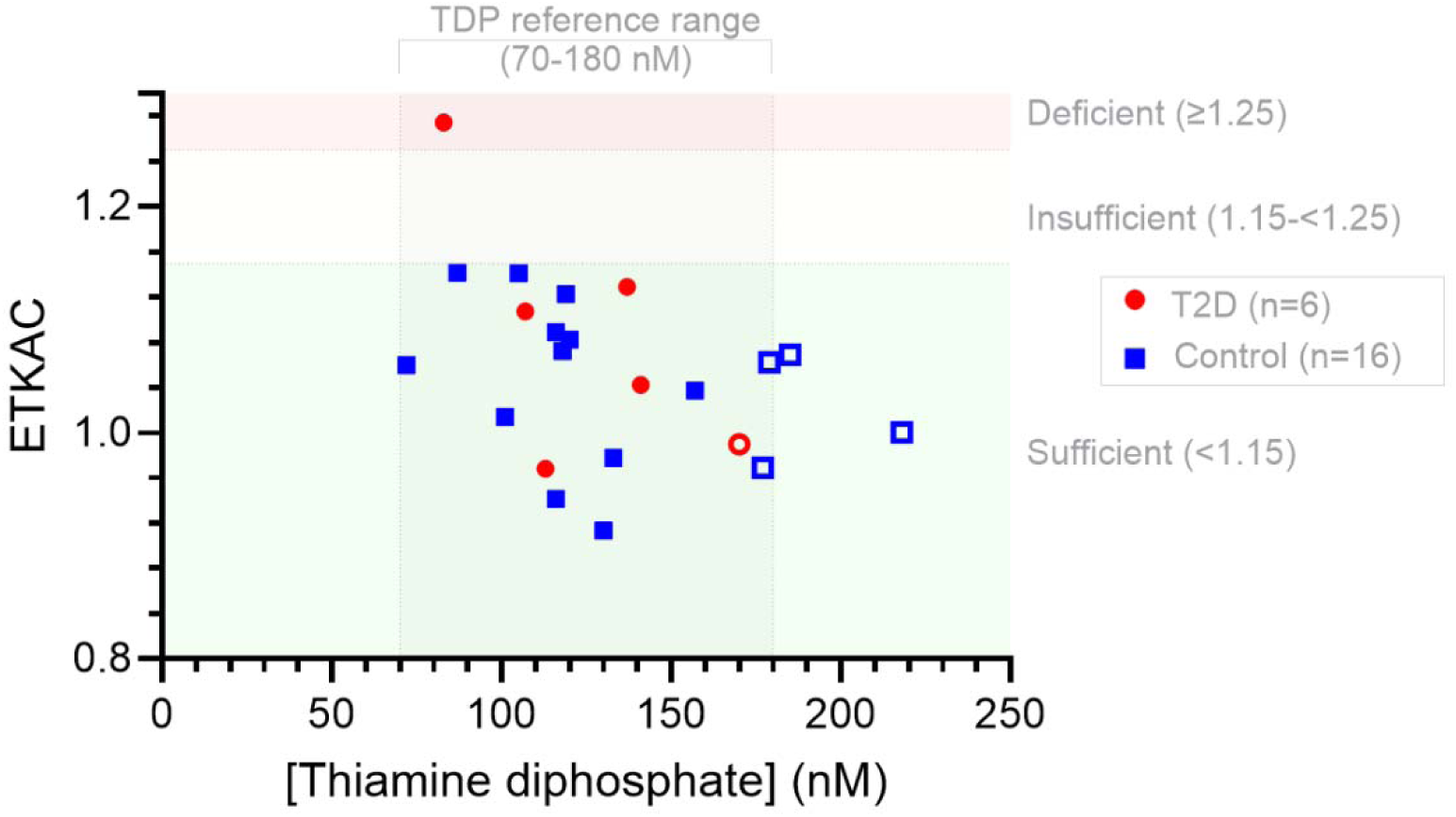
Relationship between erythrocyte transketolase activity (ETKA) ratio and whole blood thiamine diphosphate (TDP) concentrations in control participants (blue squares, n=16) and individuals with T2D (red circles, n=6). The shaded vertical region indicates the reference range for TDP (70-180 nM). Horizontal shading denotes ETKA thresholds for thiamine status: sufficient (<1.15), insufficient (1.15 - <1.25), and deficient (≥1.25). Open symbols represent participants reporting use of a thiamine-containing dietary supplement (0.8-25 mg/day)

### Erythrocyte transketolase activity varied independently of circulating thiamine diphosphate

Basal and TDP-supplemented activities were similar for most individuals, indicating minimal activation with exogenous TDP and suggesting that transketolase is largely saturated under physiological conditions. Despite this apparent overall functional sufficiency, basal TKT activity spanned approximately 6-fold across participants (Fig. 2), highlighting substantial inter-individual variability in enzyme capacity that was not explained by circulating TDP concentration. Of note, the one participant exhibiting a marked increase in activity upon TDP-supplementation already had relatively high basal activity (∼0.47 pmol/min/μg Hb, circled, Fig. 2), despite a relatively low circulating TDP concentration (83 nM). This pattern indicates substantial enzyme capacity, but incomplete coenzyme saturation.

**Fig. 2.**
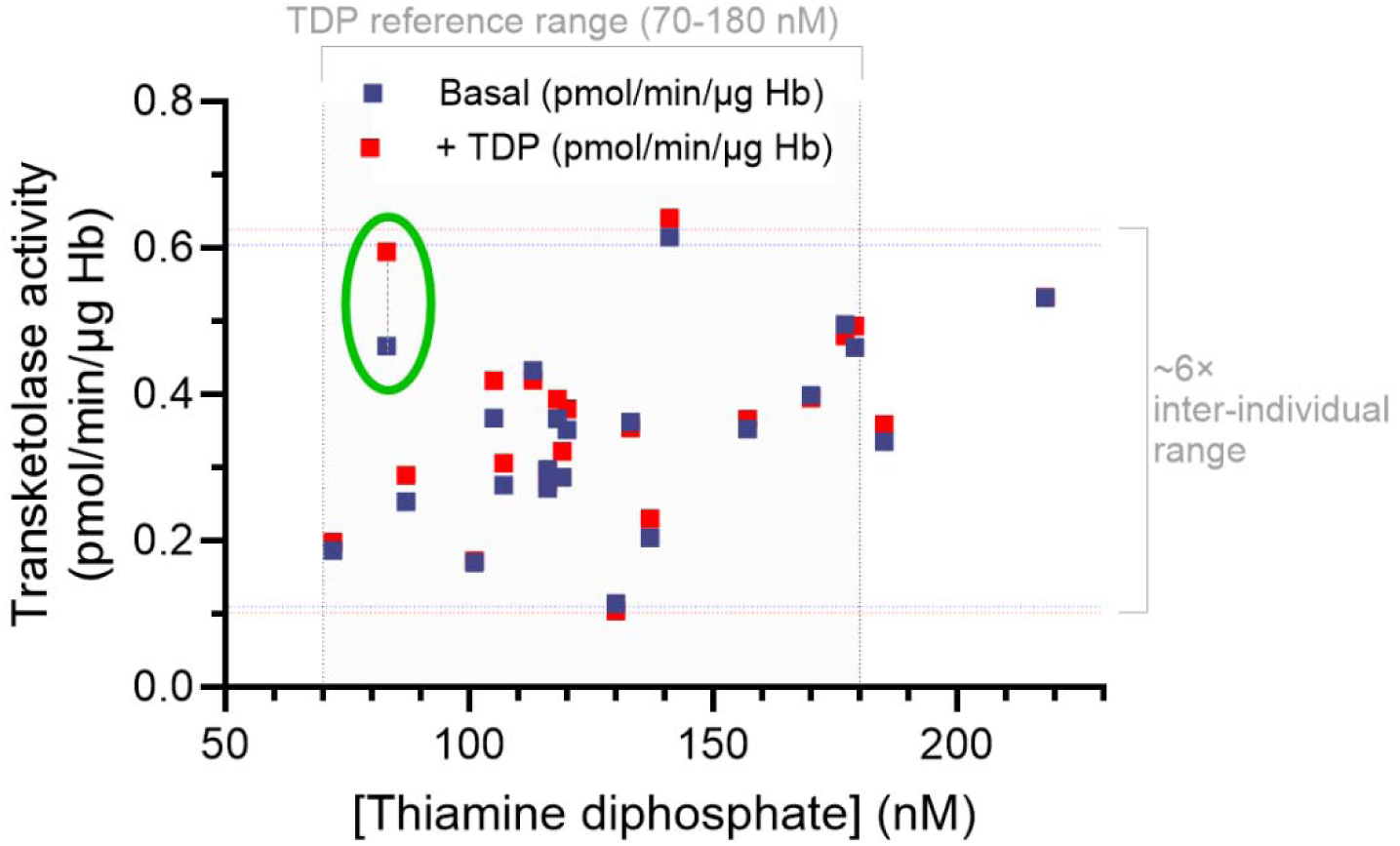
Relationship between erythrocyte transketolase specific activity and whole blood thiamine diphosphate (TDP) concentrations. Basal and TDP-supplemented activities are shown for each participant. The shaded region indicates the reference range for TDP (70-180 nM). The circled participant had an ETKAC ≥ 1.25.

### Relationship between TDP concentration and erythrocyte TKT activity differed by T2D status

When data from control and T2D participants were considered separately, TKT activity increased with whole blood TDP concentration for control participants under both basal and TDP-supplemented conditions (R^2^ = 0.54 – 0.61, Fig. 3a). The relationships were nearly identical under basal and TDP-supplemented conditions (interaction *p*=0.869), indicating that the association between circulating TDP and TKT activity persisted after ex vivo TDP supplementation. Substantial inter-individual variability remained, suggesting that TDP availability alone does not account for differences in TKT activity. The relationship was also retained when TKT activity was normalized to hemoglobin (pmol/min/μg Hb rather than pmol/min), although the association was slightly weaker.

**Fig. 3.**
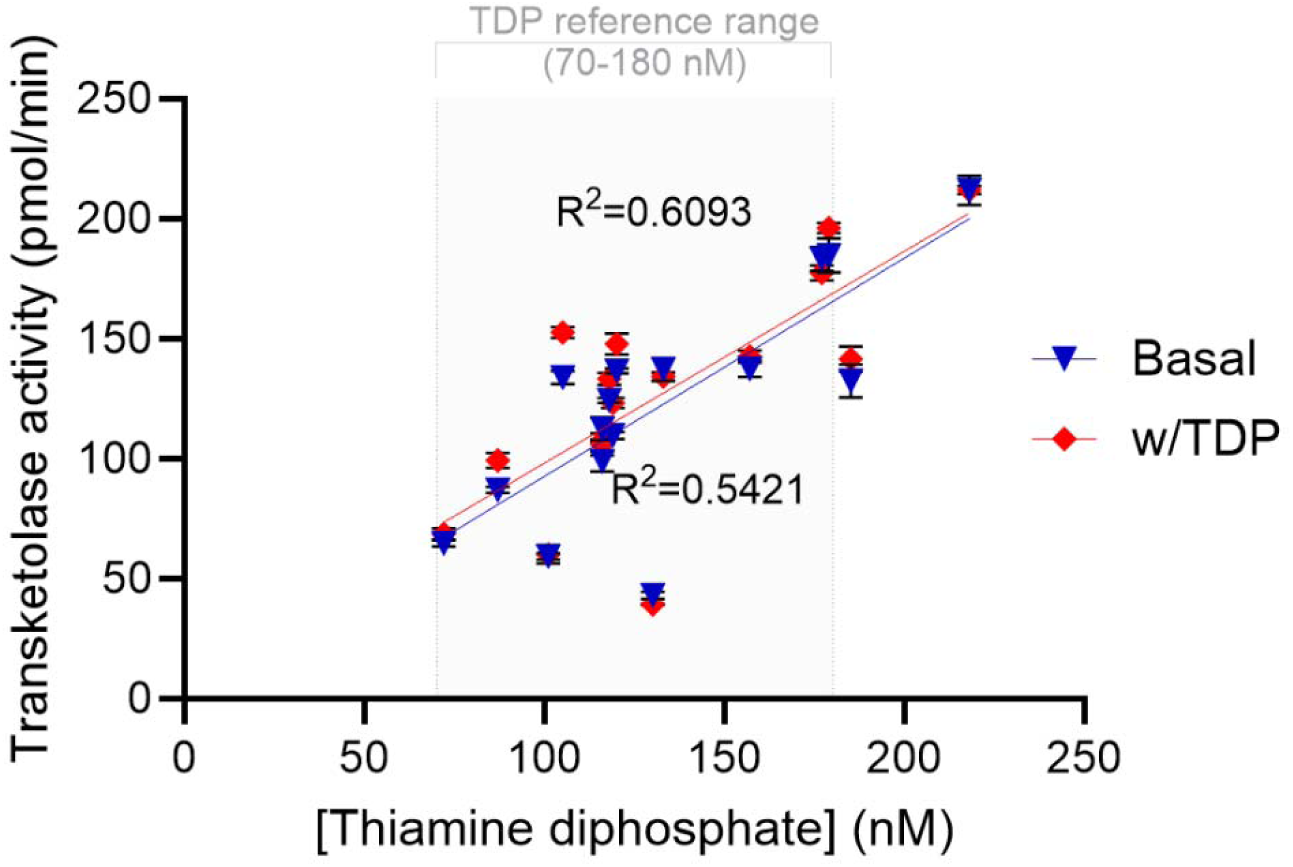
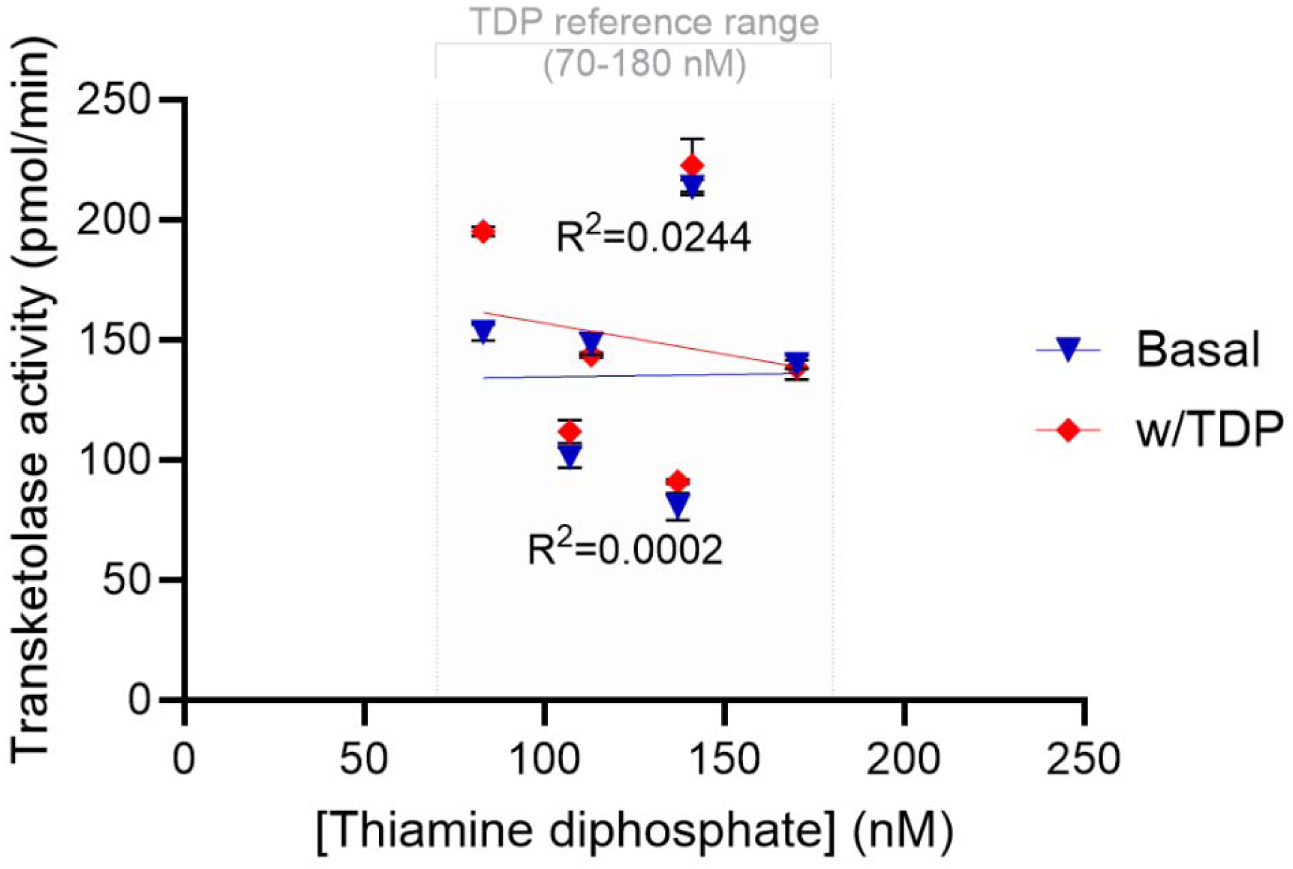
Relationship between erythrocyte transketolase activity and whole blood thiamine diphosphate (TDP) concentrations separately for A.) control and B.) T2D participants. Basal (blue triangles) and TDP-supplemented (red diamonds) TKT activities are shown with linear regression lines. The shaded region indicates the whole-blood TDP reference range. Other plot elements are described in Fig. 2.

In contrast to control participants, no relationship was observed between whole-blood TDP concentration and TKT activity in individuals with T2D (R^2^ ≈ 0; Fig. 3b). Both basal and TDP-supplemented activities showed no apparent relationship with TDP concentration, suggesting that factors beyond TDP availability contribute to inter-individual variation in TKT activity in T2D. Given the small number of T2D participants, this observation warrants further investigation.

### TDP concentrations were associated with glycemic measures in T2D but not controls

In control participants, fasting blood glucose showed little relationship with whole blood TDP concentrations. In contrast, T2D participants demonstrated a strong positive association between TDP and fasting glucose (R^2^=0.88) (Fig. 4a). Within T2D participants, the association between TDP and fasting glucose remained after adjustment for age (β=1.029 mg/dL glucose per nM TDP, *p*=0.048), whereas age was not independently associated with fasting glucose (*p*=0.786, overall model *R*^2^=0.885). A similar group-dependent relationship was observed for HbA1c and slopes again differed between T2D and control participants (interaction *p*=0.0198), although the within-T2D relationship was weaker (*R*^2^=0.62, Fig. 4b). In contrast, neither basal TKT activity nor ETKAC demonstrated corresponding associations with fasting glucose or HbA1c (Fig. S2 and S3), suggesting discordance between circulating TDP concentrations and functional TKT-based measures in relation to glycemic status.

**Fig. 4.**
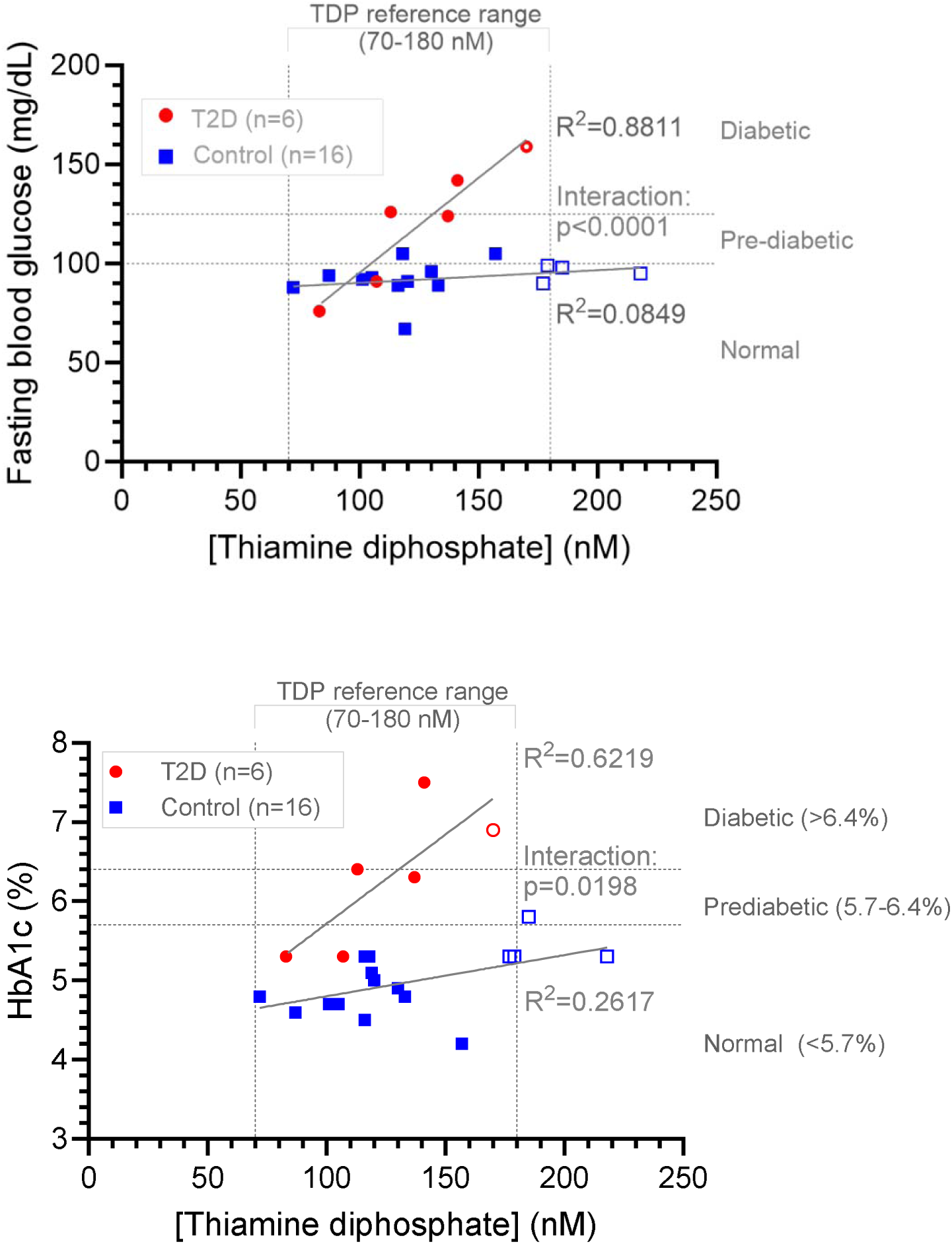
a.) Fasting blood glucose (mg/dL) and b.) hemoglobin A1c (%) relative to whole blood thiamine diphosphate concentrations (nM) in control participants (n=16, blue squares) and participants with diagnosed type 2 diabetes (n=6, red circles). Open symbols represent participants reporting use of a thiamine-containing dietary supplement (0.8-25 mg/day). The relationship between TDP and glycemic measures differed by group, with regression slopes differing significantly between control and T2D participants for fasting glucose (*p*<0.0001) and HbA1c (*p*=0.0198). Both relationships were more strongly positive in participants with T2D than in controls.

### Magnesium availability modulates transketolase activity independent of TDP supplementation

To evaluate whether added Mg^2+^ influences TKT activity, erythrocyte lysates from a subset of participants were assayed with and without exogenous magnesium. Both TDP and Mg^2+^ contribute to the formation and stabilization of active TKT dimers, and low magnesium concentrations can constrain TKT activity, independent of thiamine availability.

Across participants, addition of Mg2+ significantly increased basal TKT activity (paired t-test p=0.0048, mean increase 16.7 pmol/min), with individual increases ranging from 5.3% to 28.2% (Fig. 5a). TDP-supplemented activity was also significantly increased by Mg^2+^ (*p*=0.0164, mean increase 14.6 pmol/min), although responses were more variable, ranging from essentially no change to a 35.2% increase. The persistence of the Mg2+ effect under TDP-supplemented conditions indicates that variation in TKT activity is not attributable to TDP availability alone.

**Fig. 5.**
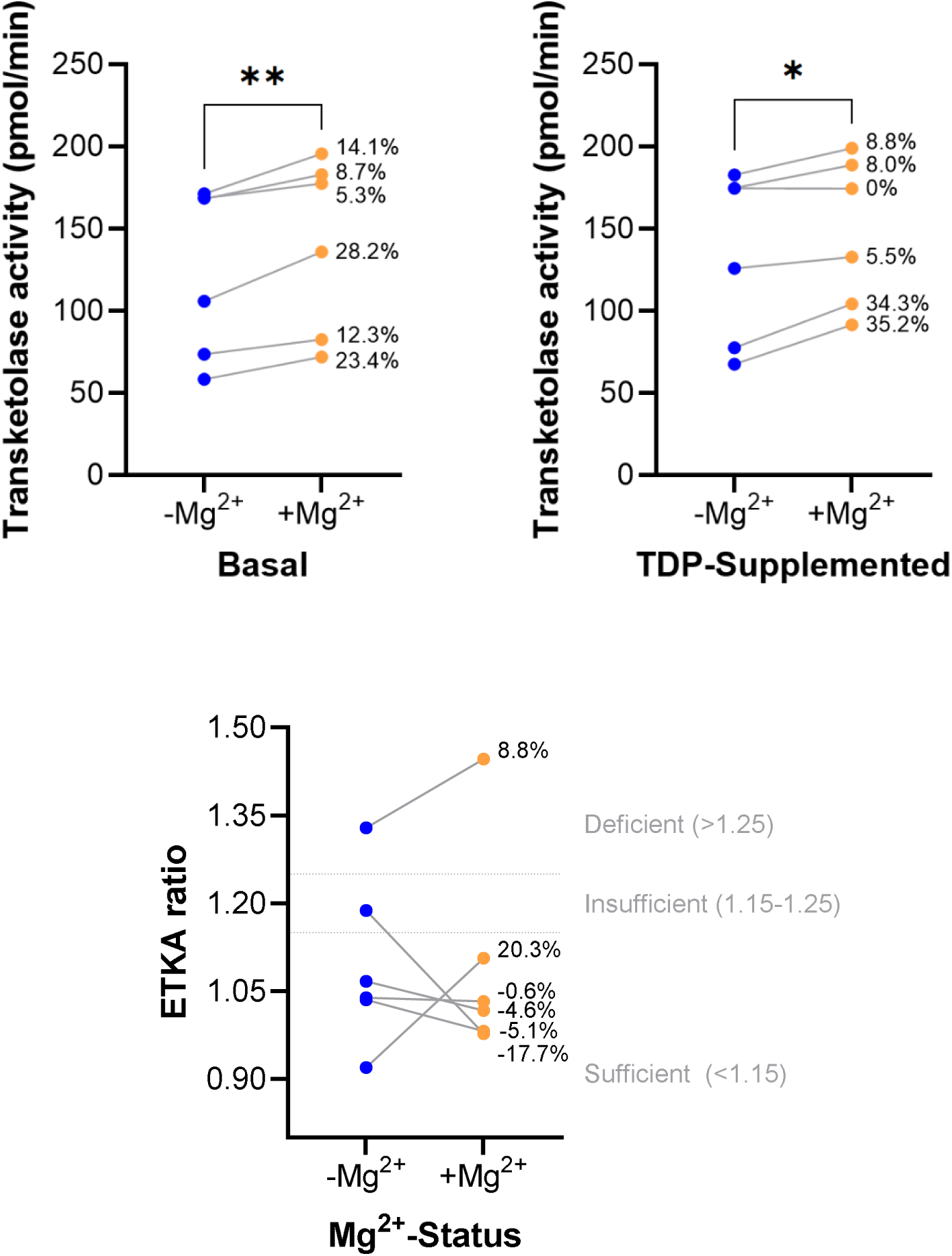
Effect of magnesium on TKT activity. Paired measurements are shown for each participant (n=6) without magnesium (-Mg^2+^) and with added magnesium (+Mg^2+^) conditions for A.) basal, B.) TDP-supplemented activity, and C.) ETKAC determinations. Each line represents an individual participant, with labels indicating the percent change following Mg^2+^ addition. Magnesium significantly increased activity overall under both conditions (paired t-test: basal *p*=0.0048, +TDP *p*=0.0164).

Normalization to hemoglobin did not materially alter the magnitude or significance of the magnesium-dependent increase in TKT activity. The enhancement of basal activity with Mg^2+^ is also consistent with the possibility that endogenous Mg^2+^ availability may constrain utilization of available TDP in some erythrocyte lysates, potentially influencing TDP utilization in vivo.

Magnesium supplementation did not consistently alter ETKAC across participants (Fig. 5c), despite increasing both basal and TDP-supplemented transketolase activities. These increases were not consistently proportional within individuals: several participants showed a greater increase in basal activity, resulting in a decreased ETKAC, while others showed a greater increase in supplemented activity, resulting in an increased ETKAC. Changes in ETKAC ranged from −17.7% to +20.3% in this small subset, demonstrating that magnesium availability can alter the resulting activation coefficient.

### NDNS dataset analyses support discordance between thiamine intake and functional status

We analyzed data from the UK NDNS to extend our analyses to a larger population and assess additional parameters unavailable in our pilot study. The NDNS dataset includes ETKAC measurements and estimates of dietary thiamine and magnesium intake, but not direct blood thiamine or TDP measurements. Adults aged 18-65 years with both ETKAC measurements and dietary survey results were included (n=3,212, Fig. 6.)

**Fig. 6.**
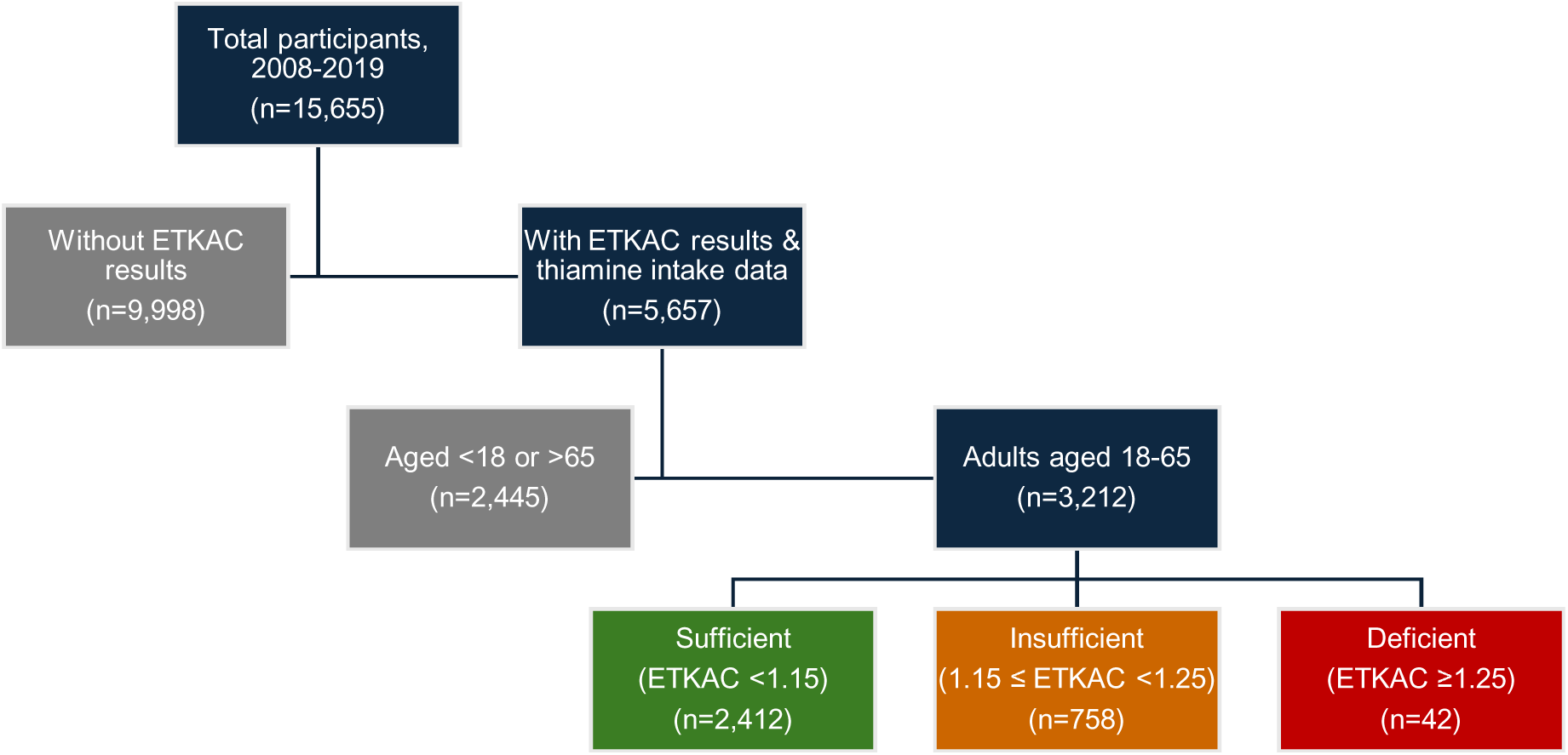
Flow diagram of NDNS participants (unweighted data) included in ETKAC analyses and stratified by age and ETKAC status. Sufficient thiamine status was defined as ETKAC<1.15, insufficient status as 1.15 ≤ ETKAC < 1.25, and deficient status as ≥ 1.25.

Among these participants, 23.6% and 1.3% were classified as functionally thiamine insufficient and deficient, respectively, despite generally adequate reported thiamine intakes. As there were relatively few individuals classified as deficient (ETKAC ≥ 1.25), data from participants with ETKAC≥ 1.15 was subsequently combined into a single non-sufficient category for further analyses. Participants classified as non-sufficient had lower magnesium intake and dietary thiamine intake (both absolute and energy adjusted) than sufficient participants and were modestly younger (Table 3). As reported by others^36^, a smaller percentage of White participants were categorized as non-sufficient than other ethnicities, most notably Black participants. However, the small subgroup sizes limit further interpretation. By contrast, the total energy intake, total sugar intake, BMI, HbA1c%, and fasting plasma glucose did not differ meaningfully between groups.

**Table 3.** Unweighted descriptive characteristics.

|  | **Sufficient\*** | **Non-sufficient\*** | **N available** | **p-value** |
| --- | --- | --- | --- | --- |
| N | 2,412 | 800 |  | - |
| Age, years | 46 (20) | 43 (21.5) | 3,212 | <0.0001 |
| BMI (kg/m<sup>2</sup>) | 26.5 (6.9) | 26.4 (6.8) | 3,114 | 0.975 |
| HbA1c (%) | 5.4 (0.5) | 5.4 (0.5) | 2,781 | 0.891 |
| Fasting blood glucose (mM) | 5.1 (0.7) | 5.1 (0.7) | 2,818 | 0.165 |
| Magnesium intake (mg/day) | 257 (123) | 244 (121) | 3,212 | <0.0001 |
| Total thiamine intake (mg/day) | 1.49 (0.79) | 1.38 (0.66) | 3,212 | <0.0001 |
| Total thiamine density (mg/1000 kcal) | 0.84 (0.40) | 0.77 (0.33) | 3,212 | <0.0001 |
| Energy intake, kcal/day | 1781 (731) | 1768 (751) | 3,212 | 0.799 |
| Total sugar intake, g/day | 86.88 (53) | 87.12 (54.75) | 3,212 | 0.938 |
| Female, n (%) | 1,426 (76.8%) | 432 (23.2%) | 3,212 | 0.011 |
| Ethnicity, n (%) |  |  |  | <0.001 |
| Asian | 71 (61.2%) | 45 (38.8%) | 116 |  |
| Black | 29 (50.9%) | 28 (49.1%) | 57 |  |
| Mixed | 20 (62.5%) | 12 (37.5%) | 32 |  |
| Other | 28 (68.3%) | 13 (31.7%) | 41 |  |
| White | 2,264 (76.3%) | 702 (23.7%) | 2,966 |  |

Continuous values are median (IQR) unless otherwise indicated. Non-sufficient functional thiamine status is defined as ETKAC ≥ 1.15. *p*-values were generated using Mann-Whitney/Wilcoxon rank-sum tests for continuous variables and Pearson chi^2^ for categorical variables. *Sample sizes vary among parameters according to data availability, as listed in the column labeled N available.

### Total dietary thiamine intake and functional status

Total thiamine intake, including supplements, was categorized by mg/day increments to facilitate interpretation (Table S1). Median ETKAC values remained within the functionally sufficient range across all intake categories, decreasing only modestly from 1.12 for participants consuming <1 mg/day to 1.09 among those consuming ≥3 mg/day. Nevertheless, the prevalence of ETKAC-defined non-sufficiency decreased from 31.4% to 17.0% with increasing thiamine intake (Fig. 7, Table S1, Pearson χ^2^(3) = 40.6, p<0.001), although substantial overlap remained between intake groups. Higher intake was associated with progressively lower predicted probabilities until approximately 2-3 mg/day, beyond which additional benefit was not evident (Fig. 7). Despite intakes ≥ 3 mg/day, 17.0% of participants remained functionally non-sufficient.

**Fig. 7.**
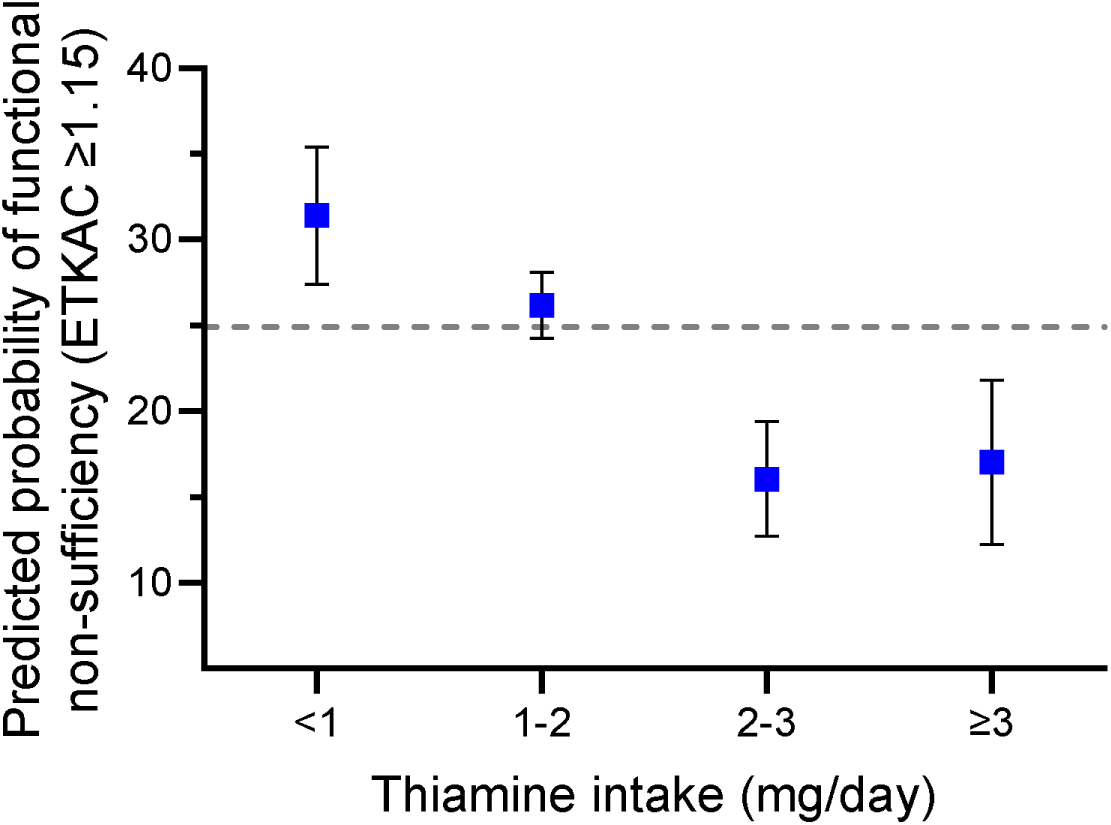
Predicted probability of functional thiamine non-sufficiency (ETKAC ≥1.15) by total thiamine intake (mg/day) in adults aged 18-65 (n=3,212). Points represent model-based estimates with 95% confidence intervals. The dashed line represents the overall predicted probability of functional non-sufficiency in the study population.

### Effect of thiamine supplements on functional thiamine status

Participants reporting the use of thiamine-containing supplements (n=369) exhibited a substantially lower prevalence of functional thiamine non-sufficiency than non-users (n=2,843) (15.7% vs 26.1%, Pearson χ^2^ =18.82, p<0.0001). However, supplemental thiamine dose was not associated with ETKAC status, suggesting a threshold rather than a dose-dependent relationship. Participants consuming ≤1.5 mg/day supplemental thiamine, consistent with typical multivitamin formulations, had a predicted probability of non-sufficiency of 16.7% compared with 26.1% among nonusers (Table S2). Higher supplemental intakes (>1.5 mg/day) were not associated with further reductions in the predicted probability of non-sufficiency (14.4% -vs-16.7%, p=0.552), consistent with a threshold effect.

### Thiamine dietary intake normalized to kcal and functional status (thiamine density)

When thiamine intake was normalized to energy intake (mg thiamine per 1000 kcal), the prevalence of ETKAC-defined non-sufficiency decreased from 38.0% among participants consuming <0.5 mg/1000 kcal to 18.0% among those consuming ≥1.0 mg/1000 kcal (Fig. 8, Table S3, Pearson χ^2^(3) = 46.8, p<0.001). Higher thiamine density was associated with progressively lower predicted probabilities. Participants consuming ≥1.0 mg/1000 kcal had significantly lower probabilities of functional non-sufficiency than all lower intake categories, although nearly one-fifth remained functionally non-sufficient.

**Fig. 8.**
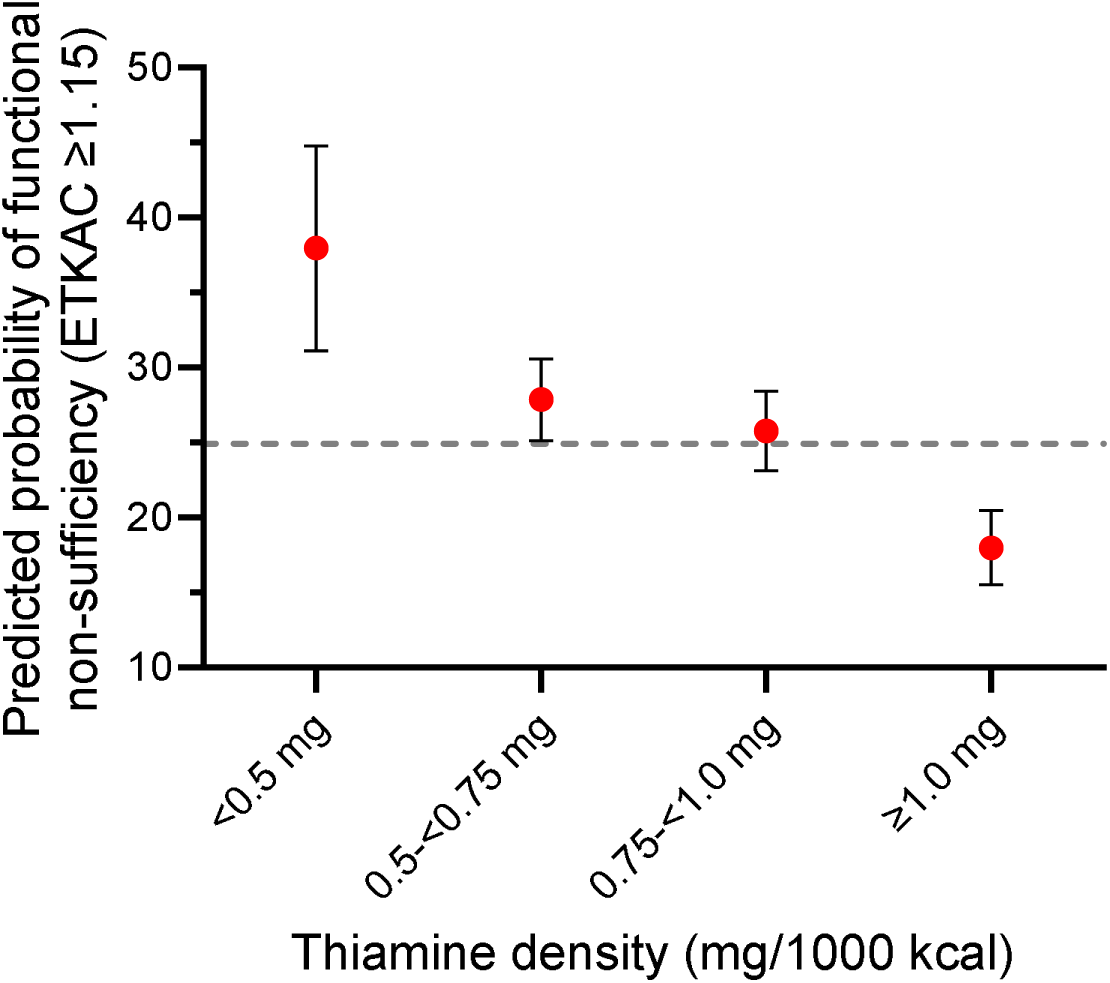
Predicted probability of functional thiamine non-sufficiency (ETKAC ≥1.15) by calorie-normalized thiamine intake (mg/1000 kcal) in adults aged 18-65 (n=3,212). Points represent model-based estimates with 95% confidence intervals. The dashed line represents the overall predicted probability of functional non-sufficiency in the study population.

### Multivariable predictors of functional thiamine non-sufficiency

To determine whether these associations were independent of demographic and metabolic factors, we fit a multivariable logistic regression model. Thiamine dietary density, age, sex, magnesium intake, and ethnicity were independently associated with functional thiamine non-sufficiency, whereas BMI and fasting blood glucose were not (Table 4).

**Table 4.** Multivariable predictors of ETKAC-defined thiamine non-sufficiency.

| Predictor | Adjusted OR (95% CI) | *p* | Overall *p* |
| --- | --- | --- | --- |
| *Thiamine density* |  |  | **<0.001** |
| <0.5 mg/1000 kcal | Reference |  |  |
| 0.5-<0.75 mg/1000 kcal | 0.602 (0.423-0.856) | **0.005** |  |
| 0.75 mg-<1.0 mg/1000 kcal | 0.530 (0.371-0.756) | **<0.001** |  |
| >=1.0 mg/1000 kcal | 0.366 (0.253-0.530) | **<0.001** |  |
| Magnesium (per 100 mg/day increase) | 0.905 (0.821-0.998) | **0.045** |  |
| Age (per 10 year increase) | 0.903 (0.841-0.969) | **0.005** |  |
| Sex (Reference: Male) | 0.825 (0.684-0.995) | **0.044** |  |
| BMI | 1.003 (0.986-1.020) | 0.752 |  |
| *Ethnicity* |  |  |  |
| White | Reference |  | **<0.001** |
| Black or Black British | 2.878 (1.637-5.060) | **<0.001** |  |
| Asian or Asian British | 2.021 (1.322-3.087) | **0.001** |  |
| Mixed Ethnic group | 1.989 (0.913-4.331) | 0.083 |  |
| Any other group | 1.617 (0.784-3.337) | 0.193 |  |
| *Fasting blood glucose* |  |  | 0.680 |
| <5.6 mM | Reference |  |  |
| 5.6-<7.0 mM | 1.107 (0.860-1.425) | 0.430 |  |
| >=7.0 mM | 1.128 (0.709-1.793) | 0.611 |  |

Adjusted odds ratios from a multivariable logistic regression model (n=2,741 complete records).
Reference categories were <0.5 mg/1000 kcal dietary thiamine density, fasting glucose <5.6 mM, male sex, and White ethnicity. Overall P values represent omnibus Wald tests for categorical predictors.

Higher thiamine density was independently associated with progressively lower odds of functional thiamine non-sufficiency, with adjusted odds ratios decreasing from 0.602 to 0.366 across increasing intake categories (Table 4). Higher magnesium intake was also independently associated with lower odds of functional non-sufficiency (OR 0.905 per 100 mg/day increase, 95% CI 0.821-0.998, *p*=0.045). There was no association of fasting blood glucose category with the adjusted odds of functional non-sufficiency. Each additional decade of age was associated with a 9.7% reduction in odds, and female sex remained independently associated with lower odds despite identical median ETKAC values between men and women (1.11). Ethnicity was strongly associated with non-sufficiency, with both Black and Asian participants having higher adjusted odds of functional thiamine non-sufficiency compared with White participants. These findings are consistent with recent analyses of the NDNS dataset demonstrating ethnic differences in ETKAC, independent of dietary intake and demographic variables.^36^ Associations of thiamine density and magnesium intake with functional non-sufficiency remained after adjustment for fasting blood glucose, BMI, and demographic characteristics. Further testing using likelihood-ratios indicated that thiamine density made a substantial independent contribution to the multivariable model, with its removal producing the largest reduction in model fit (LR χ^2^(3) = 33.66, *p*<0.001), followed by ethnicity (LR χ^2^(4) = 25.85, *p*<0.001) and age (LR χ^2^(1) = 7.93, *p*=0.0049) (Table S4). Magnesium intake and sex also made significant, but smaller contributions to model fit (Table S4.)

### Ethnicity and functional status

ETKAC distributions varied by ethnicity, with higher ETKAC values and a greater prevalence of ETKAC-defined non-sufficiency among Asian and Black participants (Table S5). These associations persisted after adjustment for dietary and metabolic covariates (Table 4). Without circulating TDP concentrations or the absolute basal and TDP-supplemented specific activities available in the NDNS, it is not possible to determine whether these differences reflect thiamine availability, differences in TKT activation, or other population-associated determinants of the ETKAC response. These findings warrant caution in assuming that differences in ETKAC-defined status necessarily reflect differences in thiamine availability alone.

### Magnesium intake and functional status

Magnesium intake was associated with functional thiamine status in univariate analyses, with the prevalence of ETKAC-defined non-sufficiency declining from 28.6% among participants consuming <200 mg/day to 21.6% among those consuming 300 - <400 mg/day (Table S6, Fig. S4, Pearson χ^2^(3) = 11.1, *p*=0.011), with little additional reduction at higher intakes. In the multivariable model, higher magnesium intake remained independently associated with lower odds of functional non-sufficiency (OR 0.905 per 100 mg/day increase, *p*=0.045), although its contribution to model fit was smaller than that of dietary thiamine density (Tables 4 and S4). There was no evidence that magnesium intake modified the association between dietary thiamine density and functional thiamine status (interaction χ^2^ =7.03, df=9, *p*=0.634, Table S7), supporting independent rather than interactive associations with ETKAC-defined status.

### Impact of diabetes status

Consistent with our pilot study, self-reported diabetes status was not associated with functional non-sufficiency (29.0% vs 25.8% in participants with and without diabetes, respectively). Pearson χ^2^(1) = 0.69, *p*=0.41, Table S8). The prevalence of ETKAC-defined thiamine non-sufficiency did not differ significantly according to self-reported diabetes status, fasting blood glucose category, or HbA1c category (Tables S9 and S10)). However, biochemical classification additionally allowed participants to be stratified into normal, prediabetic, and diabetic glycemic categories (Table S8).

## Discussion

In our study population (n=24), all participants with available TDP measurements (n=22) had whole blood TDP concentrations within or above the reference range (70-180 nM), and all but one participant had ETKAC values consistent with thiamine sufficiency (ETKAC <1.15) (Fig. 1). Nevertheless, one participant demonstrated an ETKAC consistent with functional thiamine deficiency (ETKAC ≥1.25) despite a circulating TDP concentration within the reference range (83 nM). Although near the lower end of the reference range, this concentration would not ordinarily be classified as deficient on the basis of circulating TDP alone. Possible explanations include reduced TKT affinity, limited magnesium availability, or altered enzyme stability requiring higher TDP concentrations to maintain the active dimeric form.^49–53^ It is important to note that reference ranges for whole blood TDP do not represent established diagnostic cutoffs for thiamine deficiency.^31,38,54,55^

Beyond this finding of discordance between circulating TDP and functional thiamine status, ETKAC ratio outputs can mask potentially important absolute values of TKT activity.^38,54^ We found that basal and TDP-supplemented TKT activity varied as much as six-fold among participants. These differences in absolute TKT activity were not explained by circulating TDP concentrations alone (Fig. 2). TDP concentrations and TKT activity were more closely associated in controls than T2D participants (Fig. 3), but the low sample numbers preclude extensive interpretation. Low TKT activity without elevated activation by TDP has been suggested to be due to reduced apoenzyme abundance, post-translational modifications, and low affinity variants.^56 30,57^

Such variation in absolute TKT activity may be clinically important. Elevated TKT expression and activity have been associated with poor prognosis in various cancers.^58,59^ The TKT-driven non-oxidative PPP contributes substantially to ribose-5-phosphate production, supporting the nucleotide requirements for rapidly proliferating tumor cells.^60^ Carbon recycling through TKT can also support repeated oxidative PPP flux and NADPH generation, thereby enhancing antioxidant capacity in proliferating cells.^59^ Conversely, reduced TKT activity has been observed in diabetes,^56^ diabetic neuropathy,^30^ alcohol use disorder,^49^ and Alzheimer’s Disease.^61^ Lower affinity of transketolase for TDP has been noted in Wernicke-Korsakoff syndrome and has been suggested to leave affected individuals more susceptible to thiamine deficiency.^62^ The opposing scenarios of TKT ranging from overactivity in cancer to underactivity in many other disease states suggest that greater understanding of absolute TKT values is needed.

We also noted activation of the basal and TDP-supplemented TKT activity with the addition of magnesium to erythrocyte lysates (Fig. 5), although the magnitude of activation varied among participants and consequently differentially affected their ETKAC values. Mg^2+^ facilitates the formation and stabilization of active TKT dimers and is particularly important at lower TDP concentrations; at higher levels of TDP, active dimers can form with less dependence on Mg^2+^.^63^

We observed similar Mg^2+^-dependent enhancement of TKT activity previously in fish liver extracts.^34^ Assays carried out without the addition of Mg^2+^ reflect the combined endogenous availability of TDP and Mg^2+^, whereas Mg^2+^ supplementation reduces the apparent dependence of TKT on TDP by lowering its *K_m_*. Although adding Mg^2+^ can therefore help isolate Mg^2+^ responsiveness, it also overrides variation attributable to endogenous Mg^2+^ and may provide a less physiological measure of functional thiamine status. We suggest a paired approach with and without added Mg^2+^ to ETKAC assays or including magnesium measurements on washed erythrocytes on the same samples.^34,35^

Consistent with a potential role for Mg^2+^ in TKT function in vivo, higher dietary magnesium intake was weakly associated with greater functional thiamine sufficiency in the NDNS cohort (Table 3). However, dietary intake and serum magnesium are imperfect proxies for intracellular or erythrocyte magnesium availability, with serum magnesium representing a small fraction of total body stores.^64^ Consistent with our ex vivo pilot study results, previous studies likewise found that intravenous thiamine administered with magnesium increased erythrocyte TKT activity more than thiamine alone^65^ and that low serum magnesium (<0.75 mM) was associated with a weaker relationship between basal TKT activity and erythrocyte TDP concentration.^66^ These results collectively highlight the importance of understanding a subject’s magnesium levels alongside the interpretation of functional thiamine availability. The cause of low TKT activities or non-sufficient ETKAC values could be probed by using higher concentrations of supplemental TDP or supplemental magnesium in the assay,^49,52,67–69^ which could be incorporated into the workflow for follow up of samples.

As low circulating TDP concentrations were not consistently associated with elevated ETKAC or reduced TKT activities in our pilot cohort, circulating TDP levels within the reference range appear sufficient for substantial TKT saturation in most individuals. However, coupled with the deficient ETKAC result described above, these findings suggest that circulating TDP concentration alone may not reliably predict functional thiamine status.

In the NDNS cohort, after adjustment for age, sex, ethnicity, BMI, fasting plasma glucose, and magnesium intake, dietary thiamine density (mg/1000 kcal) remained one of the strongest independent predictors of functional thiamine status (Table 4). The recommended daily allowance (RDA) for thiamine in the U.S. is 1.2 mg for men, 1.1 mg for women, and 1.4 mg during pregnancy and lactation. In the U.K., it is 0.8 mg for women and 1.0 mg for men, aged 19-64.^1^ Given the role of thiamine in glucose handling, an RDA based on the caloric intake of 0.5 mg/1000 kcal has also been proposed.^1,26,43^ This approach is consistent with the concept that thiamine requirements should be considered relative to metabolic substrate load.^70^ In the present study, the probability of functional thiamine non-sufficiency plateaued at approximately 2-3 mg/day, while progressively decreasing with increasing thiamine densities above 0.5 mg/1000 kcal. Although total energy intake was nearly identical between functionally sufficient and non-sufficient participants (Table 3), higher thiamine density remained independently associated with functional sufficiency, suggesting that thiamine exposure relative to energy intake, rather than total energy intake, was more strongly related to functional thiamine status. These findings suggest that thiamine intakes associated with biochemical sufficiency may exceed those required to prevent overt deficiency. Even at the highest levels (≥3 mg/day or ≥1 mg/1000 kcal), ∼17% of participants remained functionally non-sufficient. Our results suggest that ETKAC-defined functional thiamine non-sufficiency is relatively common despite dietary intakes generally considered sufficient.

In our pilot study, we did not see differences between participants with and without T2D in whole blood TDP concentrations, absolute TKT activities, or ETKAC values. Similarly, in the NDNS cohort, ETKAC-defined non-sufficiency was not significantly associated with self-reported diabetes status, fasting glucose category, or % HbA1c category (Tables 4 and S8-S10). In patients with diabetes, plasma thiamine deficiency has gone unrecognized as TKT activity is often within a normal range.^71^ This discrepancy has been attributed in part to increased levels of thiamine transporters in erythrocytes of diabetic patients, allowing for greater uptake from the circulation.^71^ Enhanced erythrocyte uptake could therefore preserve TKT function and reduce the likelihood of elevated ETKAC despite lower circulating plasma TDP concentrations. Such compartmental differences could contribute to discordance between circulating and functional measures of thiamine status. Whole-blood TDP measurements in our pilot study cannot distinguish the erythrocyte contribution from plasma thiamine, and future studies incorporating paired plasma thiamine and erythrocyte or whole-blood TDP measurements may help resolve these relationships.

We did, however, observe an association of increased circulating TDP with increased fasting blood glucose in participants with T2D, whereas this relationship was absent in controls (Fig. 4). A similar group-dependent pattern was observed for HbA1c. One possible explanation is that higher carbohydrate intake may simultaneously increase glycemic exposure and thiamine intake, given that many staple carbohydrate-containing foods in the United States are fortified with thiamine. However, the small number of participants with T2D and absence of dietary intake records preclude evaluation of this possibility. Importantly, corresponding associations with glycemic measures were not observed for TKT activity or ETKAC in the pilot cohort, and ETKAC was unrelated to glycemic category in the larger NDNS cohort. Together, these observations further suggest that circulating TDP and TKT-based functional measures may capture different aspects of thiamine biology in relation to glycemic status.

Ethnicity was also independently associated with ETKAC-defined functional status, with higher odds of non-sufficiency among Black and Asian participants relative to White participants after adjustment for dietary thiamine density, magnesium intake, age, sex, BMI, and fasting glucose. This finding is consistent with a recent separate analysis of the NDNS reporting ethnic differences in ETKAC that persisted after adjustment for dietary and demographic factors.^36^ Notably, that analysis found that adjustment for dietary thiamine intake did not account for ethnic differences in ETKAC. However, these differences cannot necessarily be assumed to represent differences in thiamine availability or requirement itself. In the absence of circulating TDP concentrations and absolute basal and TDP-supplemented TKT activities in the NDNS, differences in ETKAC could reflect variation in thiamine availability, TKT abundance or activation, cofactor interactions, or other determinants of erythrocyte function. These findings therefore reinforce the broader conclusion of this study that ETKAC is a functional assay where biological interpretation may not be equivalent across individuals or populations.

### Limitations

Our pilot study enrolled a relatively small number of participants, limiting statistical power and broader generalization. Dietary intake data were not collected in the pilot cohort. To complement these findings, we used the NDNS dataset, which provided population-based data on ETKAC, body measures, dietary intake, and biochemical parameters beyond those obtained in our pilot study. However, differences in dietary patterns and food fortification practices between the U.K. and the U.S. may limit direct extrapolation between populations. The corresponding publicly accessible U.S. survey (CDC NHANES) provides dietary intake information but does not include measurements of thiamine levels or functional status. Dietary survey results rely on participant recall on dietary intake and nutritional content databases rather than direct measurement of thiamine or magnesium levels. The specific basal or TDP-supplemented activity values were also not available in this dataset. While diabetes status in the NDNS was based on self-reporting, the availability of measured fasting blood glucose and HbA1c values provided an independent means of assessing glycemic status. However, the NDNS did not distinguish T1D from T2D. The ethnic subgroup sample sizes in the NDNS study were also small. We note that the NDNS analyses were cross-sectional and unweighted and therefore were intended to evaluate associations within the analytical sample rather than estimate nationally representative prevalence.

## Conclusions

Circulating TDP, dietary thiamine intake, and ETKAC provide related but non-equivalent information about thiamine status. Dietary thiamine density was strongly associated with ETKAC-defined functional status in the NDNS cohort, yet substantial non-sufficiency persisted even at relatively high intakes, while our pilot study demonstrated marked inter-individual variation in absolute TKT activity that was not captured by ETKAC or circulating TDP alone.

Magnesium availability further altered TKT activity and, in some individuals, the resulting ETKAC response. Demographic and dietary factors, including ethnicity and magnesium intake, were also independently associated with ETKAC, emphasizing that functional measurements may reflect determinants beyond thiamine intake alone. Together, these findings support an integrated approach to thiamine assessment incorporating dietary exposure, circulating TDP, and functional enzyme measurements, including absolute TKT activity where feasible. Such an approach may better distinguish inadequate thiamine availability from variation in cofactor utilization and intrinsic enzyme capacity than any single measure alone.

## Acknowledgements

We gratefully acknowledge the laboratory staff at United Health Services and Terri Peters, RN, UHS Research Program Manager, for their logistical support and drawing and submitting blood samples for analysis. We thank Lindsey Moon for her contributions to the in-house laboratory work. Funding for this study was in part through a Binghamton University Health Sciences Transdisciplinary Areas of Excellence Award. We thank the participants in our pilot study and staff and participants in the NDNS surveys.

## Authorship Contributions

KE and NC secured funding and approvals for this study. NC conducted interviews, carried out body-mass measurements, POC assays, and all interactions with human subjects. PW and ML completed in-house laboratory work. KE wrote the original draft of the manuscript, supervised the laboratory work, processed TKT data, and designed the NDNS study and carried out NDNS analyses. All authors reviewed and approved the content of this manuscript.

## Data availability

The NDNS data described in the manuscript are publicly available from the United Kingdom Data Service. Data generated in the pilot study that support the findings of this study are available from the corresponding author upon reasonable request.

## Notes

### Competing Interest Statement

The authors have declared no competing interest.

### Author Declarations

IRB of Binghamton University gave ethical approval for this work

