## Supporting information for "Circulating thiamine diphosphate, dietary intake, and transketolase function are non-equivalent measures of thiamine status"

*Corresponding author

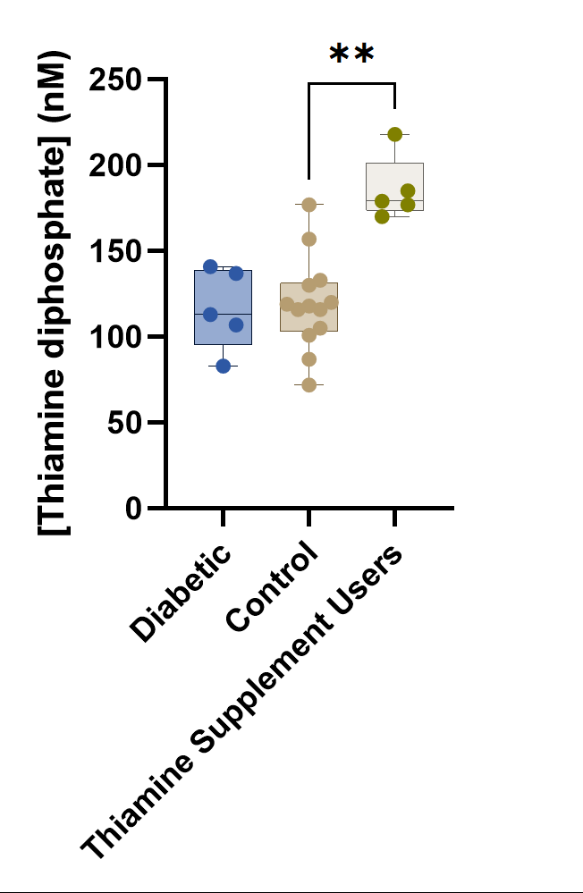

**Fig. S1.** TDP concentrations in control, T2D, and thiamine supplement users. TDP concentrations were higher among thiamine supplement users (including both control and T2D participants).

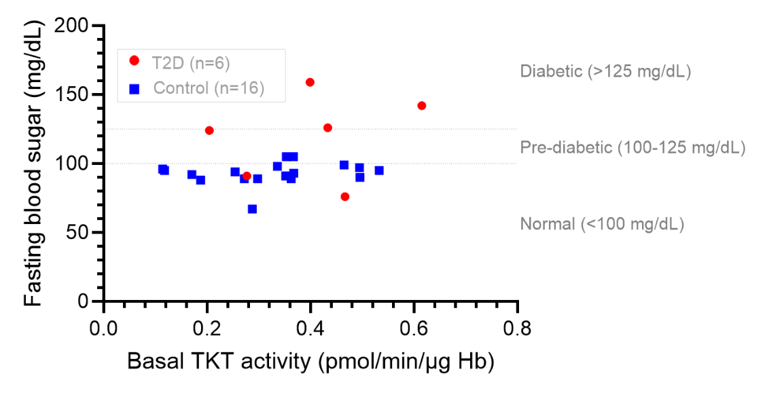

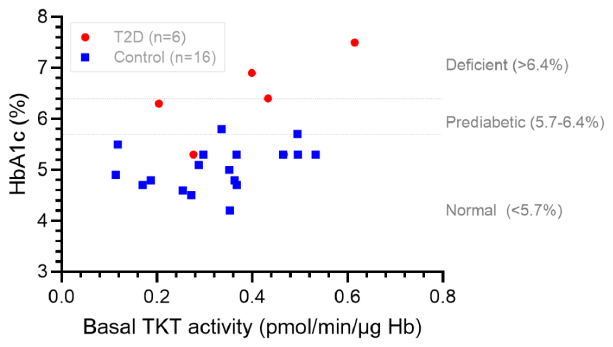

**Fig. S2.** Relationships between basal TKT activity and a.) fasting blood glucose and b.) HbA1c in control participants and participants with T2D.

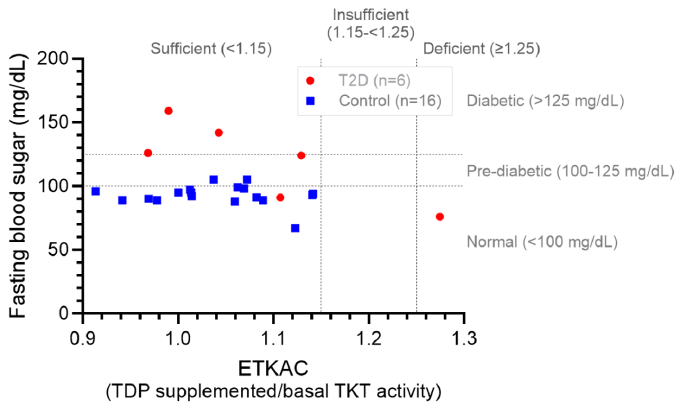

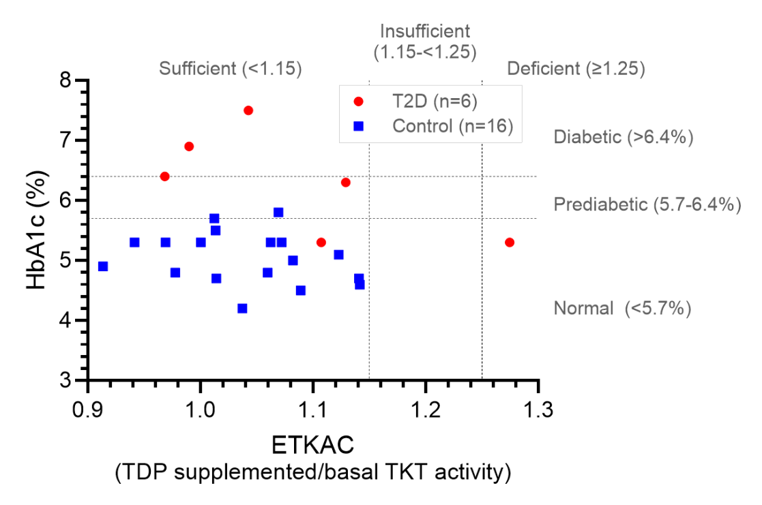

**Fig. S3.** Relationships between ETKAC and a.) fasting blood glucose and b.) HbA1c in control participants and participants with T2D.

**Table S1**. ETKAC status as a function of thiamine intake, including supplements

|  |  | **ETKAC status** | | | | |
| --- | --- | --- | --- | --- | --- | --- |
| **Thiamine intake** | **Median ETKAC (IQR)** | **Sufficient** | **Insufficient** | **Deficient** | **Non-sufficient** | **Total** |
| <1.0 mg/day | 1.12 (1.08-1.15) | 356 (68.59%) | 148 (28.52%) | 15 (2.89%) | 163 (31.41%) | 519 (100%) |
| 1 - <2 mg/day | 1.11 (1.08-1.15) | 1,474 (73.81%) | 498 (24.94%) | 25 (1.25%) | 523 (26.19%) | 1,997 (100%) |
| 2 - <3 mg/day | 1.10 (1.06-1.13) | 387 (83.95%) | 72 (15.62%) | 2 (0.43%) | 74 (16.05%) | 461 (100%) |
| ≥3 mg/day | 1.09 (1.05-1.13) | 195 (82.98%) | 40 (17.02%) | 0 (0%) | 40 (17.02%) | 235 (100%) |
| *Total* | 1.11 (1.08-1.14) | 2,412 (75.09%) | 758 (23.60%) | 42 (1.31%) | 800 (24.91%) | 3,212 (100%) |

*Values shown are participant counts with percentages of sufficient, insufficient, deficient, or cumulatively non-sufficient respectively.*

**Table S2.** ETKAC status as a function of thiamine supplement dose

|  |  | **ETKAC status** | | | | |
| --- | --- | --- | --- | --- | --- | --- |
| **Thiamine Supplement dose** | **Median ETKAC (IQR)** | **Sufficient** | **Insufficient** | **Deficient** | **Non-sufficient** | **Total** |
| 0 mg | 1.13 (1.10-1.17) | 2,101 (73.90%) | 700 (24.62%) | 42 (1.48%) | 742 (26.10%) | 2,843 (100%) |
| ≤1.5 mg | 1.11 (1.08-1.15) | 180 (83.33%) | 36 (16.67%) | 0 (0%) | 36 (16.67%) | 216 (100%) |
| >1.5 mg | 1.11 (1.08-1.15) | 131 (85.62%) | 22 (14.38%) | 0 (0%) | 22 (14.38%) | 153 (100%) |
| *Total* | 1.11 (1.08-1.14) | 2,412 (75.09%) | 758 (23.60%) | 42 (1.31%) | 800 (24.91%) | 3,212 (100%) |

**Table S3.** ETKAC status as a function of thiamine intake per 1000 kcal

|  |  | **ETKAC status** | | | | |
| --- | --- | --- | --- | --- | --- | --- |
| **Thiamine intake** | **Median ETKAC (IQR)** | **Sufficient** | **Insufficient** | **Deficient** | **Non-sufficient** | **Total** |
| <0.5 mg/1000 kcal | 1.13 (1.10-1.17) | 121 (62.05%) | 66 (33.85%) | 8 (4.10%) | 74 (37.95%) | 195 (100%) |
| 0.5-<0.75 mg/1000 kcal | 1.11 (1.08-1.15) | 744 (72.16%) | 272 (26.38%) | 15 (1.45%) | 287 (27.84%) | 1,031 (100%) |
| 0.75-<1.0 mg/1000 kcal | 1.11 (1.08-1.15) | 781 (74.24%) | 257 (24.43%) | 14 (1.33%) | 271 (25.76%) | 1,052 (100%) |
| ≥1.0 mg/1000 kcal | 1.10 (1.06-1.13) | 766 (82.01%) | 163 (17.45%) | 5 (0.54%) | 168 (17.99%) | 934 (100%) |
| *Total* | 1.11 (1.08-1.14) | 2,412 (75.09%) | 758 (23.60%) | 42 (1.31%) | 800 (24.91%) | 3,212 (100%) |

*Values shown are participant counts with percentages of sufficient, insufficient, deficient, or cumulatively non-sufficient respectively.*

**Table S4.** Contributions of individual parameters to multivariable model fit

| Predictor | LR χ^2^ | *df* | Overall *p* |
| --- | --- | --- | --- |
| *Thiamine density* | 33.66 | 3 | **<0.001** |
| *Ethnicity* | 25.85 | 4 | **<0.001** |
| *Fasting blood glucose* | 0.77 | 2 | 0.6815 |
| *Magnesium intake (per 100 mg/day intake increase)* | 4.08 | 1 | **0.0433** |
| *Sex* | 4.05 | 1 | **0.0441** |
| *Age (per 10 year increase)* | 7.93 | 1 | **0.0049** |
| *BMI* | 1.71 | 1 | 0.1911 |

Likelihood-ratio tests compared the full multivariable logistic regression model with otherwise identical models omitting each predictor individually. *p* values are derived from likelihood-ratio tests and therefore may differ slightly from Wald test *p* values reported in Table 4.

**Table S5.** ETKAC and ETKAC-defined functional thiamine status according to ethnicity in NDNS participants

| **Ethnicity** | **n** | **ETKAC median (IQR)** | **Non-sufficiency, n (%)** | **Adjusted OR (95% CI)** |
| --- | --- | --- | --- | --- |
| White | 2,966 | 1.11 (1.07-1.14) | 702 (23.7%) | Reference |
| Asian | 116 | 1.12 (1.09-1.18) | 45 (38.8%) | 2.021 (1.322-3.087) |
| Black | 57 | 1.14 (1.10-1.17) | 28 (49.1%) | 2.878 (1.637-5.060) |
| Mixed | 32 | 1.13 (1.08-1.17) | 12 (37.5%) | 1.989 (0.913-4.331) |
| Other | 41 | 1.13 (1.09-1.15) | 13 (31.7%) | 1.617 (0.784-3.337) |

Descriptive statistics include all participants with available ETKAC and ethnicity data (n=3,212). Adjusted Ors are derived from the complete-case multivariable model (n=2,741; Table 4).

**Table S6.** ETKAC status as a function of magnesium intake

|  |  | **ETKAC status** | | | | |
| --- | --- | --- | --- | --- | --- | --- |
| **Magnesium intake** | **Median ETKAC (IQR)** | **Sufficient** | **Insufficient** | **Deficient** | **Non-sufficient** | **Total** |
| <200 mg | 1.12 (1.08-1.15) | 569 (71.39%) | 210 (26.35%) | 18 (2.26%) | 228 (28.61%) | 797 (100%) |
| 200-<300 mg | 1.11 (1.08-1.15) | 1,038 (74.95%) | 335 (24.19%) | 12 (0.87%) | 347 (25.05%) | 1,385 (100%) |
| 300-<400 mg | 1.10 (1.07-1.14) | 595 (78.39%) | 154 (20.29%) | 10 (1.33%) | 164 (21.61%) | 759 (100%) |
| ≥400 mg | 1.10 (1.07-1.14) | 210 (77.49%) | 59 (21.77%) | 2 (0.74%) | 61 (22.51%) | 271 (100%) |
| *Total* | 1.11 (1.08-1.14) | 2,412 (75.09%) | 758 (23.60%) | 42 (1.31%) | 800 (24.91%) | 3,212 (100%) |

Pairwise comparisons of model-predicted probabilities of functional insufficiency (ETKAC ≥ 1.15) demonstrated that intake of 300 – <400 mg magnesium/day differed significantly from <200 mg/day (p = 0.002, Fig. S4).

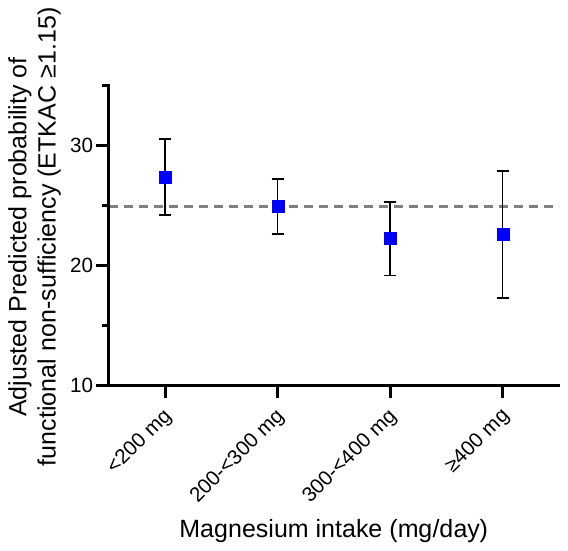

**Fig. S4.** Adjusted predicted probability of functional thiamine non-sufficiency (ETKAC ≥1.15) by dietary magnesium intake (mg) in adults aged 18-65 (n=3,212). Points represent model-based estimates with 95% confidence intervals. The dashed line represents the overall predicted probability of non-sufficiency in the study population.

**Table S7.** Thiamine magnesium effects on non-sufficiency status

| Thiamine density (adjusted for magnesium) | Higher thiamine intake associated with lower odds of non-sufficiency | Independent association |
| --- | --- | --- |
| Magnesium intake (adjusted for thiamine) | Higher magnesium intake associated with lower odds of non-sufficiency | Independent association |
| Thiamine density x magnesium interaction | Pearson χ^2^ =7.03, df=9, p=0.634 | No evidence that magnesium modifies the thiamine-density association |

**Table S8.** Diabetes status counts by self-reported versus biochemical glycemic categories

|  | **Self-reported** | **FBG** | **HbA1c** |
| --- | --- | --- | --- |
| Non-diabetic/Normal | 2,817 (726/25.8%) | 2,261 (581/25.7%) | 2,038 (532/26.1%) |
| Pre-diabetic | N/A | 446 (120/26.9%) | 631 (158/25.0%) |
| Diabetic | 131 (38/29.0%) | 111 (31/27.9%) | 112 (35/31.3%) |
| Total | 2,948 | 2,818 | 2,781 |
| *p* | 0.409 | 0.774 | 0.385 |

Numbers in parentheses are the # of non-sufficient individuals in each category, followed by the percentage.

**Table S9.** ETKAC status delineated by fasting blood glucose clinical glycemic categories

|  |  | **ETKAC status** | | | | |
| --- | --- | --- | --- | --- | --- | --- |
| **Fasting blood glucose categorization** | **Median ETKAC (IQR)** | **Sufficient** | **Insufficient** | **Deficient** | **Non-Sufficient** | **Total** |
| Normal | 1.11 (1.08-1.15) | 1,680 (74.30%) | 555 (24.55%) | 26 (1.15%) | 581 (25.70%) | 2,261 (100%) |
| Prediabetes | 1.11 (1.08-1.15) | 326 (73.09%) | 107 (23.99%) | 13 (2.91%) | 120 (26.91%) | 446 (100%) |
| Diabetes | 1.12 (1.08-1.15) | 80 (72.07%) | 29 (26.13%) | 2 (1.80%) | 31 (27.93%) | 111 (100%) |
| *Total* | 1.11 (1.08-1.15) | 2,086 (74.02%) | 691 (24.52%) | 41 (1.45%) | 732 (25.98%) | 2,818 (100%) |

**Table S10.** ETKAC status delineated by HbA1c % clinical glycemic categories

|  |  | **ETKAC status** | | | | |
| --- | --- | --- | --- | --- | --- | --- |
| **HbA1c% categorization** | **Median ETKAC (IQR)** | **Sufficient** | **Insufficient** | **Deficient** | **Non-Sufficient** | **Total** |
| Normal | 1.11 (1.08-1.15) | 1,506 (73.90%) | 507 (24.88%) | 25 (1.23%) | 532 (26.10%) | 2,038 (100%) |
| Prediabetes | 1.11 (1.07-1.15) | 473 (74.96%) | 146 (23.14%) | 12 (1.90%) | 158 (25.04%) | 631 (100%) |
| Diabetes | 1.12 (1.08-1.16) | 77 (68.75%) | 33 (29.46%) | 2 (1.79%) | 35 (31.25%) | 112 (100%) |
| *Total* | 1.11 (1.08-1.15) | 2,056 (73.93%) | 686 (24.67%) | 39 (1.40%) | 725 (26.07%) | 2,781 (100%) |
